# Artificial Intelligence in Pancreatic Intraductal Papillary Mucinous Neoplasm Imaging: A Systematic Review

**DOI:** 10.1101/2025.01.08.25320130

**Authors:** Muhammad Ibtsaam Qadir, Jackson A. Baril, Michele T. Yip-Schneider, Duane Schonlau, Thi Thanh Thoa Tran, C. Max Schmidt, Fiona R. Kolbinger

## Abstract

**Background:** Based on the Fukuoka and Kyoto international consensus guidelines, the current clinical management of intraductal papillary mucinous neoplasm (IPMN) largely depends on imaging features. While these criteria are highly sensitive in detecting high-risk IPMN, they lack specificity, resulting in surgical overtreatment. Artificial Intelligence (AI)-based medical image analysis has the potential to augment the clinical management of IPMNs by improving diagnostic accuracy.

**Methods:** Based on a systematic review of the academic literature on AI in IPMN imaging, 1041 publications were identified of which 25 published studies were included in the analysis. The studies were stratified based on prediction target, underlying data type and imaging modality, patient cohort size, and stage of clinical translation and were subsequently analyzed to identify trends and gaps in the field.

**Results:** Research on AI in IPMN imaging has been increasing in recent years. The majority of studies utilized CT imaging to train computational models. Most studies presented computational models developed on single-center datasets (n=11,44%) and included less than 250 patients (n=18,72%). Methodologically, convolutional neural network (CNN)-based algorithms were most commonly used. Thematically, most studies reported models augmenting differential diagnosis (n=9,36%) or risk stratification (n=10,40%) rather than IPMN detection (n=5,20%) or IPMN segmentation (n=2,8%).

**Conclusion:** This systematic review provides a comprehensive overview of the research landscape of AI in IPMN imaging. Computational models have potential to enhance the accurate and precise stratification of patients with IPMN. Multicenter collaboration and datasets comprising various modalities are necessary to fully utilize this potential, alongside concerted efforts towards clinical translation.

**Graphical Abstract:** 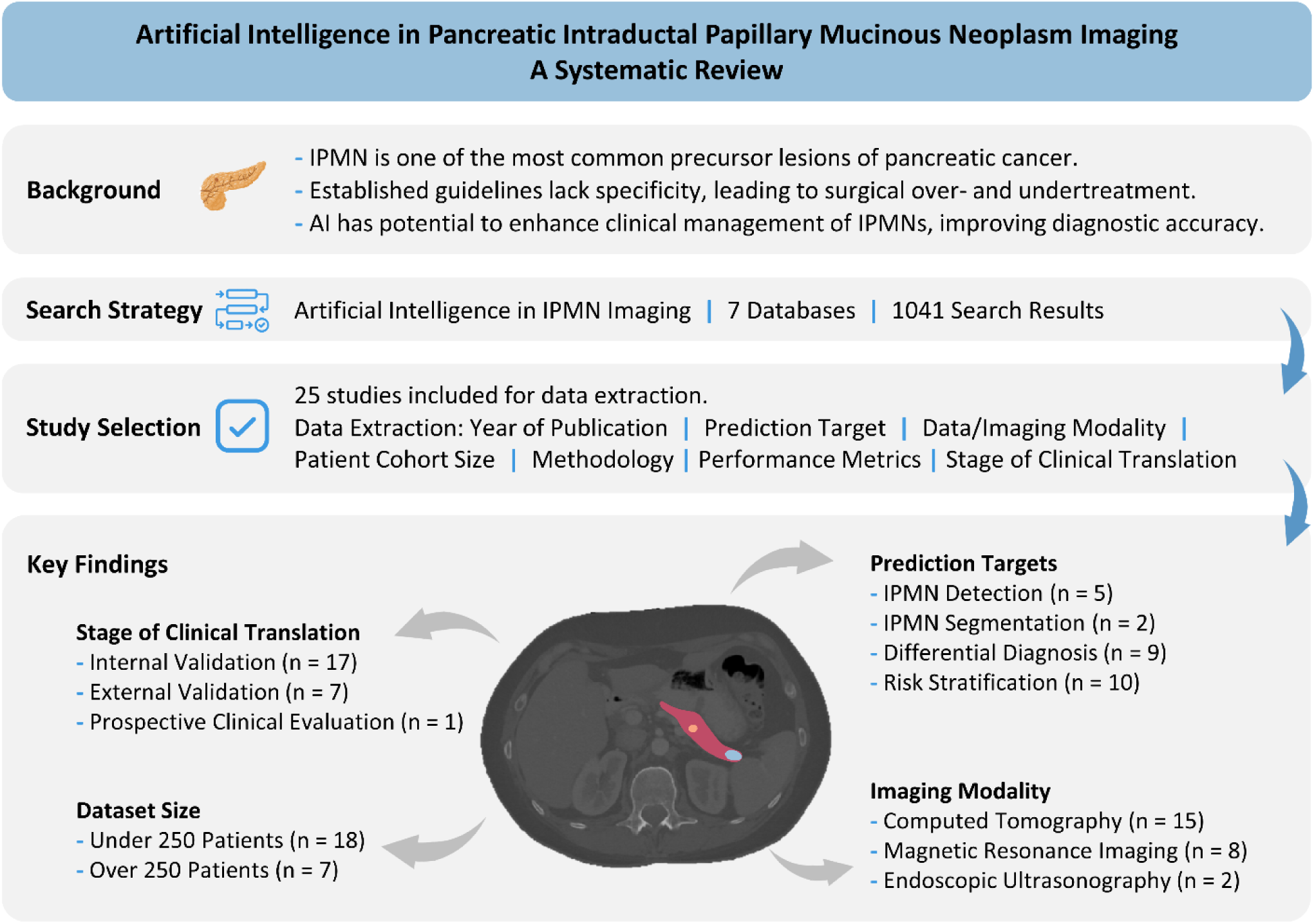

**Highlights:**

- Artificial Intelligence holds promise in the field of IPMN by augmenting IPMN detection, differentiation of different types of pancreatic cysts, stratifying malignant progression risk, and automating the analysis of IPMN imaging through computational cyst segmentation.
- The majority of studies related to AI-based analysis of IPMN imaging use single-center patient cohorts of less than 250 patients to develop and validate computational models and consider imaging as the only data modality.
- Reporting transparency of existing studies on AI in IPMN imaging is limited and there remains a scarcity of comprehensive, multimodal approaches as well as clinical translation.

## Introduction

Pancreatic cancer is one the most aggressive cancers, with a 5-year overall survival rate that remains under 15% [1,2]. Pancreatic ductal adenocarcinoma (PDAC) is the most prevalent solid tumor of the pancreas and constitutes more than 85% of all pancreatic cancer incidences [3,4]. The majority of patients receive a diagnosis at a late and incurable stage of the disease. Some pancreatic cysts, often incidentally detected on imaging [5], are potential precursors to pancreatic cancer, presenting a window of opportunity for early detection and cancer prevention. The most common type of pancreatic cyst is Intraductal papillary mucinous neoplasm (IPMN) [6]. About 10% of the general population over 50 years of age are estimated to carry IPMN [7]. The frequency of invasive carcinoma in resected branch duct (BD-)IPMN and main duct (MD-)IPMN ranges from 10 to 45% [8–10] and from 40 to 75% [10–12], respectively. In patients with BD-IPMN, the 5-year and 15-year malignant progression rates are 3.3% and 12%, respectively [13]. Of all PDACs, 8-10% are estimated to derive from IPMN [14,15].

The management of IPMN primarily depends on imaging features, cytology, and clinical variables such as CA19-9 serum levels and pancreatitis [7,16]. Patients with MD-IPMN and pancreatic duct dilation to a diameter of 10 mm or more or a nodule of at least 5 mm are generally recommended surgical resection; in patients with BD-IPMN, imaging features such as main duct dilation to a diameter of 5-9 mm, enhancing mural nodule less than 5 mm, cyst size ≥ 30 mm or a cyst growth rate ≥ 2.5 mm/year are considered indicative of higher malignant potential warranting consideration for surgical resection [10]. Patients carrying BD-IPMN without such “worrisome features” are recommended surveillance every 6 or 12 months, including imaging and a clinical examination [10]. Given the substantial risks associated with pancreatic surgery including mortality [17] and morbidity [18] such as post-pancreatectomy diabetes mellitus [19], recommendations for surgical resection are generally determined by experts who evaluate each case using all available diagnostic modalities [20]. The Fukuoka [10] and Kyoto international consensus guidelines [21] represent the gold standard for IPMN management; however, while they are sensitive, they lack specificity [22], Consequently, patient risk stratification is often not accurate, either overestimating malignant transformation risk, resulting in surgical overtreatment [23,24], or underestimating the malignant potential, resulting in surveillance of malignant IPMN (undertreatment) [25].

Artificial Intelligence (AI), particularly deep learning, can detect characteristic patterns in large medical imaging datasets such as radiological, histopathological, and endoscopic imaging, that are beyond what humans can identify [26,27]. Similarly, traditional natural language processing (NLP) models and, more recently, large language models (LLMs), facilitate the interpretation of written reports, which may support clinical decision-making and personalization of patient care [28,29]. The complex management of IPMN patients is an ideal use case for AI-based computational tools, which could contribute to clinical decision-making and risk stratification based on available data, particularly imaging as the primary non-invasive source of information about IPMN malignant potential.

This systematic review provides an overview of the scientific literature on AI in IPMN imaging. We summarize relevant clinical and technological aspects of published studies, including patient selection criteria and cohort sizes, prediction targets, data modalities, methodological characteristics, performance metrics, translational considerations, and data availability. By highlighting the research trends and gaps, this review aims to showcase the potential of computational methods in the context of the clinical management of IPMN.

## Methods

### Search Strategy

We performed this systematic review following the PRISMA (Preferred Reporting Items for Systematic Reviews and Meta-Analyses) [30] and AMSTAR (A MeaSurement Tool to Assess systematic Reviews) [31] guidelines. To cover the interdisciplinary literature on AI in IPMN imaging, our search strategy included four literature databases (PubMed, IEEE Xplore, Embase, and CINAHL [Cumulative Index to Nursing and Allied Health Literature]), and three preprint servers (ArXiv, medRxiv, and bioRxiv). Three authors with experience in medical AI research (MIQ, JAB and FRK) and a librarian focusing on biomedical literature contributed to the development of the search strategy. Medical subject headings (MeSH) terms were used to specify the search term.

PubMed, IEEE Xplore, Embase, CINAHL, and ArXiv were searched on January 31, 2024, without language restrictions, for literature published since database inception, using the following search term: ((“Pancreatic Neoplasms”[MeSH Terms]) OR (“Pancreatic Cyst”[MeSH Terms]) OR (“Pancrea* Neoplasm*”[Title/Abstract])) AND ((“Diagnostic Imaging”[MeSH Terms]) OR (diagnosis[MeSH Terms]) OR (Record*[Title/Abstract]) OR (Tomography[Title/Abstract]) OR (Radiolog*[Title/Abstract]) OR (methods[MeSH Terms]) OR (“Electronic Medical Record”[Title/Abstract]) OR (“EMR”[Title/Abstract]) OR (Report*[Title/Abstract])) AND ((“Artificial Intelligence”[MeSH Terms]) OR (“Deep Learning”[Title/Abstract]) OR (“Natural Language Processing”[MeSH Terms]) OR (“neural network*”[Title/Abstract]) OR (“Machine Learning”[Title/Abstract]) OR (AI[Title/Abstract])). The format of the search term was modified per the databases’ requirements. MedRxiv and bioRxiv were searched on January 31, 2024, using the following, shorter search term to comply with the character restrictions of the respective search tools: (“Learning” OR “Artificial Intelligence”) AND (“Pancreas” OR “Pancreatic”) AND (“Imaging” OR “EMR”).

### Study Selection

Duplicate studies were removed automatically and manually. All studies were independently screened by two raters: one physician (JAB) with three years of experience in IPMN management and research and one engineer (MIQ) with two years of experience in AI and computational medical image analysis, using the Covidence platform (Covidence systematic review software, Veritas Health Innovation, Melbourne, Australia, https://www.covidence.org/). Screening results were blinded until the completion of each reviewer’s individual screening. The screening process was carried out in two iterations: First, screening based on title and abstract, and second, full-text screening. In each iteration, conflicts between the two reviewers were resolved by independent assessment of a third reviewer (FRK).

The inclusion criteria were: (i) analysis of IPMN (and synonyms: mucinous cyst, mucinous cystic lesion) imaging, (ii) development of computational models (machine learning, deep learning, or NLP) for the analysis of medical images and/or imaging reports, (iii) imaging modality being Endoscopic Ultrasonography (EUS) or tomography, i.e. Magnetic Resonance Imaging (MRI) or Computed Tomography (CT), (iv) original research, (v) analysis of human data, and (vi) the article being in English. Non-original articles and publications (i) not analyzing any data from patients with IPMN or (ii) not developing computational models for data analysis (i.e., statistical methods, manually curated imaging features), were excluded.

### Data Extraction and Analysis

One reviewer with expertise in AI and deep learning-based medical image analysis (MIQ) extracted the following data from each included study based on a data extraction protocol (Supplementary Table 1): (i) year of publication, (ii) prediction targets, (iii) ground truth source, (iv) number of centers in the study, (v) patient selection criteria, (vi) total number of patients on which the AI model was developed, (vii) number of patients with IPMN on which the AI model was developed, (viii) number of patients in the training set, (ix) number of patients in test/validation set, (x) validation procedure, (xi) data/imaging type, (xii) computational methodology, (xiii) model performance, (xiv) stage of clinical translation, and (xv) data availability. To assess the risk of bias, two reviewers (MIQ and JAB) independently evaluated the studies using PROBAST [32] (prediction model risk of bias assessment tool), with a third reviewer (FRK) to resolve discordances. The overall risk of bias for each study was categorized as low, high, or unclear, based on the evaluations across four domains: participants, predictors, outcomes, and analysis.

We systematically categorized the studies into four use cases based on the prediction targets: (i) detection, (ii) segmentation, (iii) differential diagnosis, and (iv) risk stratification. Detection encompasses the identification of the presence of pancreatic cysts including IPMN. Segmentation refers to the precise localization of the pancreatic cyst. This use case included publications developing a segmentation method for pixel- or voxel-wise delineation of IPMN or other pancreatic cysts in the radiological images. Differential diagnosis concentrates on differentiating between types of pancreatic cysts including cysts other than IPMN. Risk stratification includes classifying the IPMN and other pancreatic cysts into IPMN with low-grade dysplasia (benign cysts) or IPMN with high-grade dysplasia (malignant cysts). To provide a comprehensive account of the previous research, we used descriptive statistics to examine the data for each category. Key findings are summarized to highlight trends and gaps in the current literature.

### Study Registration

This systematic review was prospectively registered at OSF on January 4, 2024 (https://doi.org/10.17605/OSF.IO/NVGXB) and at PROSPERO on August 13, 2024 (CRD42024575960). The protocol was not amended or changed.

## Results

### Search Results

The search strategy, when queried across all the databases and preprint servers, resulted in 1041 publications (PubMed: n = 558, Embase: n = 199, CINAHL: n = 51, IEEE Xplore: n = 45, bioRxiv: n = 137, medRxiv: n = 47, arXiv: n = 4, Figure 1). After the removal of duplicates (n = 125), 916 unique publications were screened.

**Figure 1:**
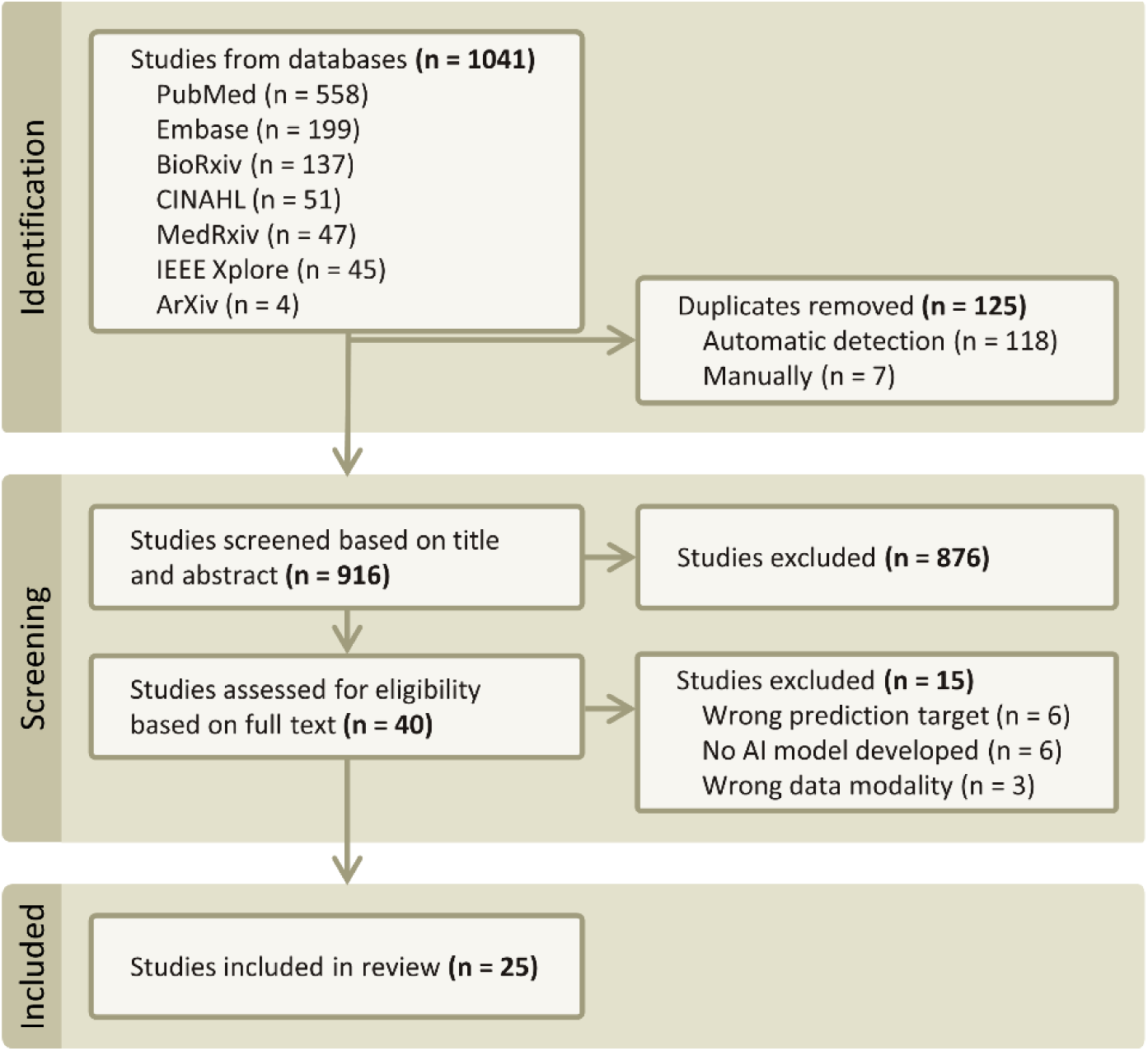
Flowchart of the systematic review and meta-analysis according to the PRISMA 2020 statement for reporting systematic reviews. Based on the systematic review of the publications on AI in IPMN imaging, 25 articles published between 2018 and 2023 were included in this analysis.

Based on title and abstract screening, 40 studies were selected for full-text screening. A total of 15 publications were excluded based on their prediction target (n = 6), data modality (n = 6) or for not developing any AI model (n = 3). Specifically, studies involving deep learning-based MRI reconstruction were excluded, as these fell outside the scope of diagnosis or risk stratification of IPMN. Additionally, studies using imaging modalities other than MRI, CT, or EUS were excluded to align with consensus guidelines for IPMN management [10,21] and American College of Radiology’s (ACR) Appropriateness Criteria for pancreatic cysts [33]. This resulted in the inclusion of 25 publications in the systematic review (Figure 1).

### Study Characteristics

Studies were categorized based on the analyzed imaging modality. The majority of publications on AI in IPMN imaging (n = 15, 60%) analyzed CT imaging, one-third of the studies (n = 8, 32%) employed MRI imaging, while only a few (n = 2, 8%) were based on EUS imaging (Figure 2).

**Figure 2:**
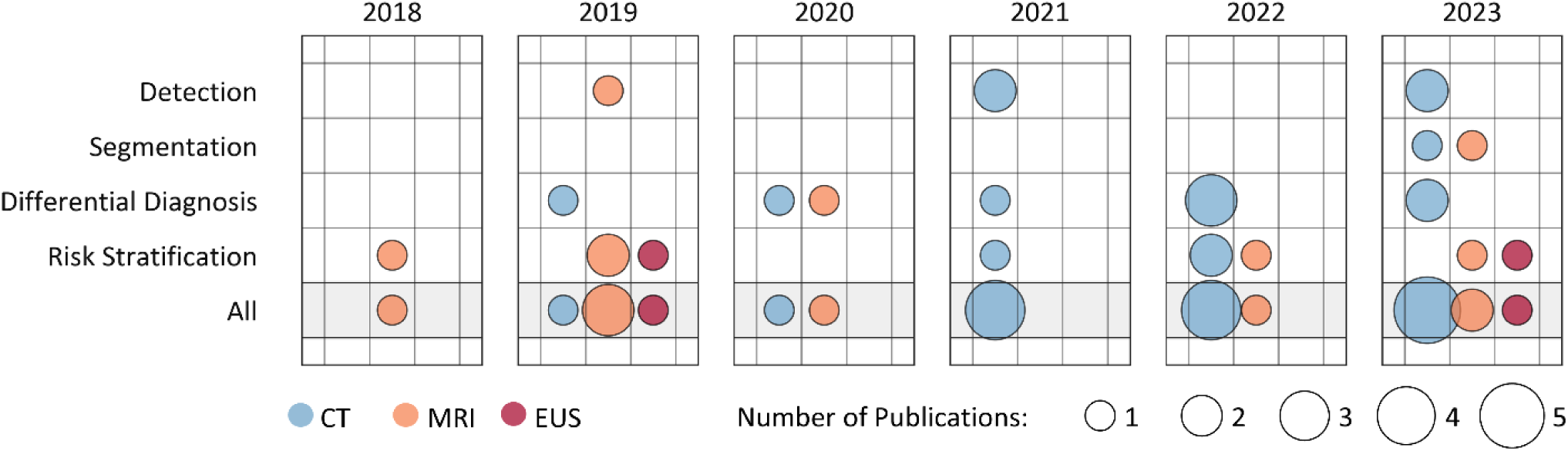
Published studies by year of publication and prediction target. The studies are further categorized by the imaging modality used. The radius of each circle is proportional to the number of publications, with colors indicating imaging modality.

We scrutinized the publication trend over time, both collectively and for each prediction target. All included studies were conducted between 2018 and 2023. The number of publications increased over time, with more than two-thirds (n = 17, 68%) of all the studies published within the last three years (2021-2023), and the highest number of studies published in 2023 (n = 8, 32%, Figure 2).

Based on the prediction target, approximately one-fourth of the included studies (n = 5, 20%) focused on detection, a couple of studies (n = 2, 8%) on segmentation, while the majority of the studies performed differential diagnosis (n = 9, 36%) and risk stratification (n = 10, 40%) of pancreatic cysts and IPMN (Figure 3).

**Figure 3:**
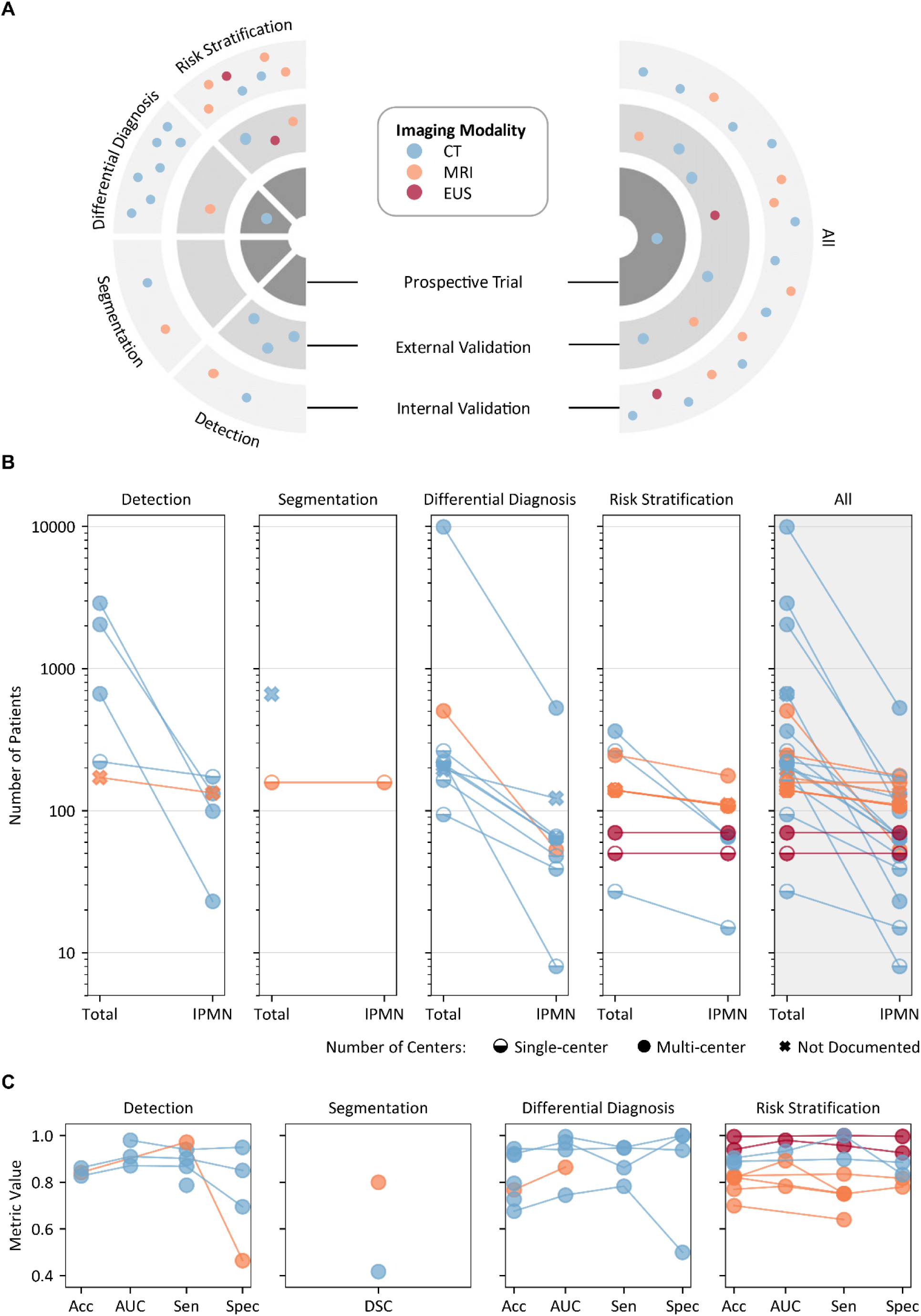
Overview of research on AI in IPMN imaging. **(A)** Distribution of publications on AI in IPMN imaging based on prediction target (IPMN/cyst detection, cyst segmentation, differential diagnosis, risk stratification) and stage of clinical translation: internal validation (monocentric studies), external validation (multicentric studies), prospective clinical evaluation. Each dot represents a single publication, with the dot color indicating the imaging modality used. **(B)** Size of the total patient cohort and the patient cohort with IPMN. Colors indicate the imaging modality used, and symbol shapes indicate the number of centers involved in the study. One study evaluating segmentation did not document the number of IPMN patients in the underlying cohort. **(C)** Performance metrics - Accuracy (Acc), Area Under the Receiver Operating Curve (AUC), Sensitivity (Sen), and Specificity (Spec) - for the proposed model across prediction targets in each study. Colors represent the imaging modalities used. For segmentation, the DSC is presented. Not all studies reported every metric.

### Current Landscape and Clinical Translation of AI in IPMN Imaging

Regarding the stage of clinical translation, over two-thirds of the studies (n = 17, 68%) conducted internal validation, 7 (28%) carried out external validation, and only one study (4%) proceeded to a prospective clinical evaluation (Figure 3 A). Nearly half of the studies (n = 11, 44%) were single-center studies, and one-third (n = 8, 32%) were multicenter studies, with most of these being two-center studies (n = 5, 20%). The remaining studies (n = 6, 24%) did not document the number of centers explicitly (Figure 3 B, Table 1).

**Table 1:**
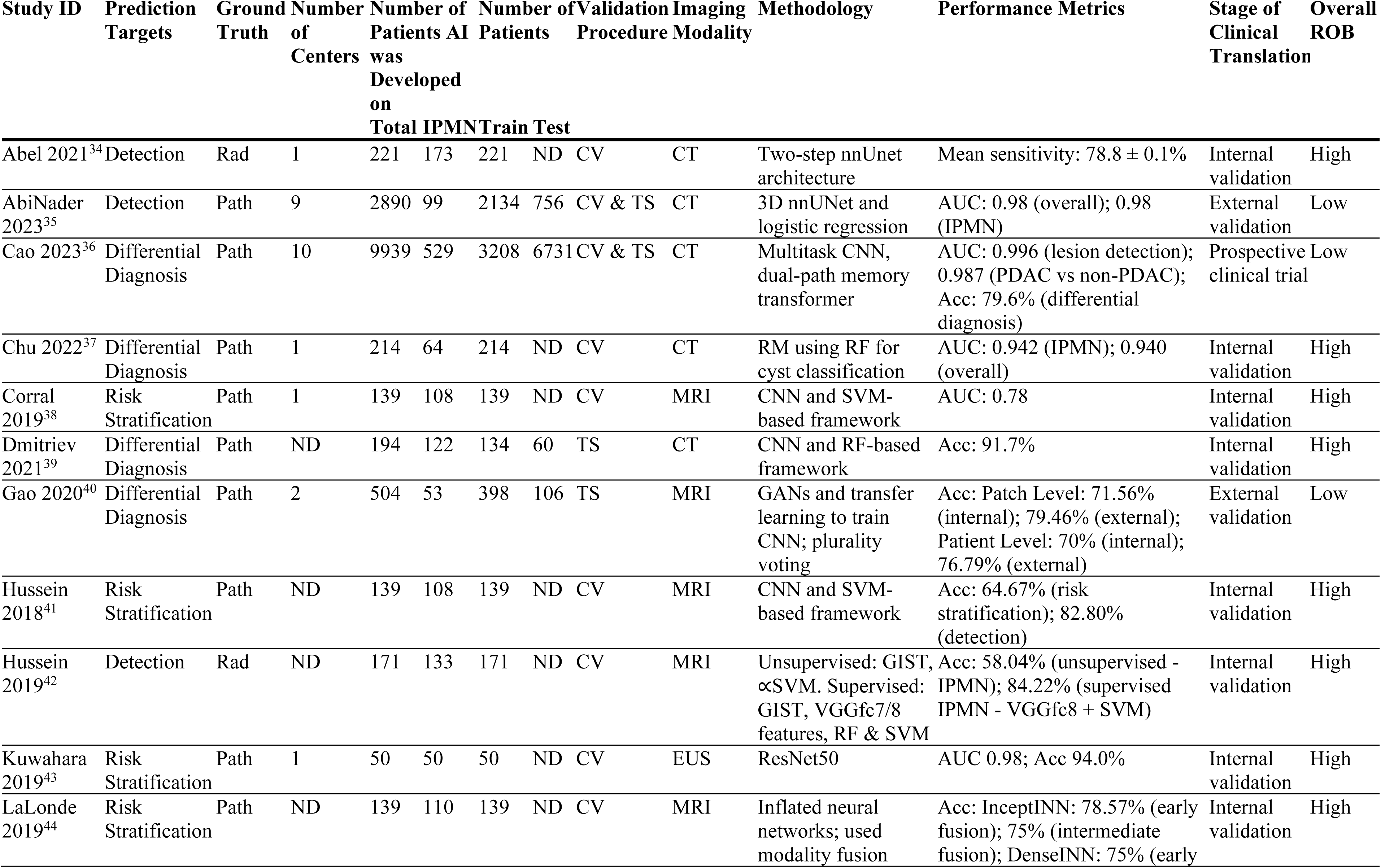
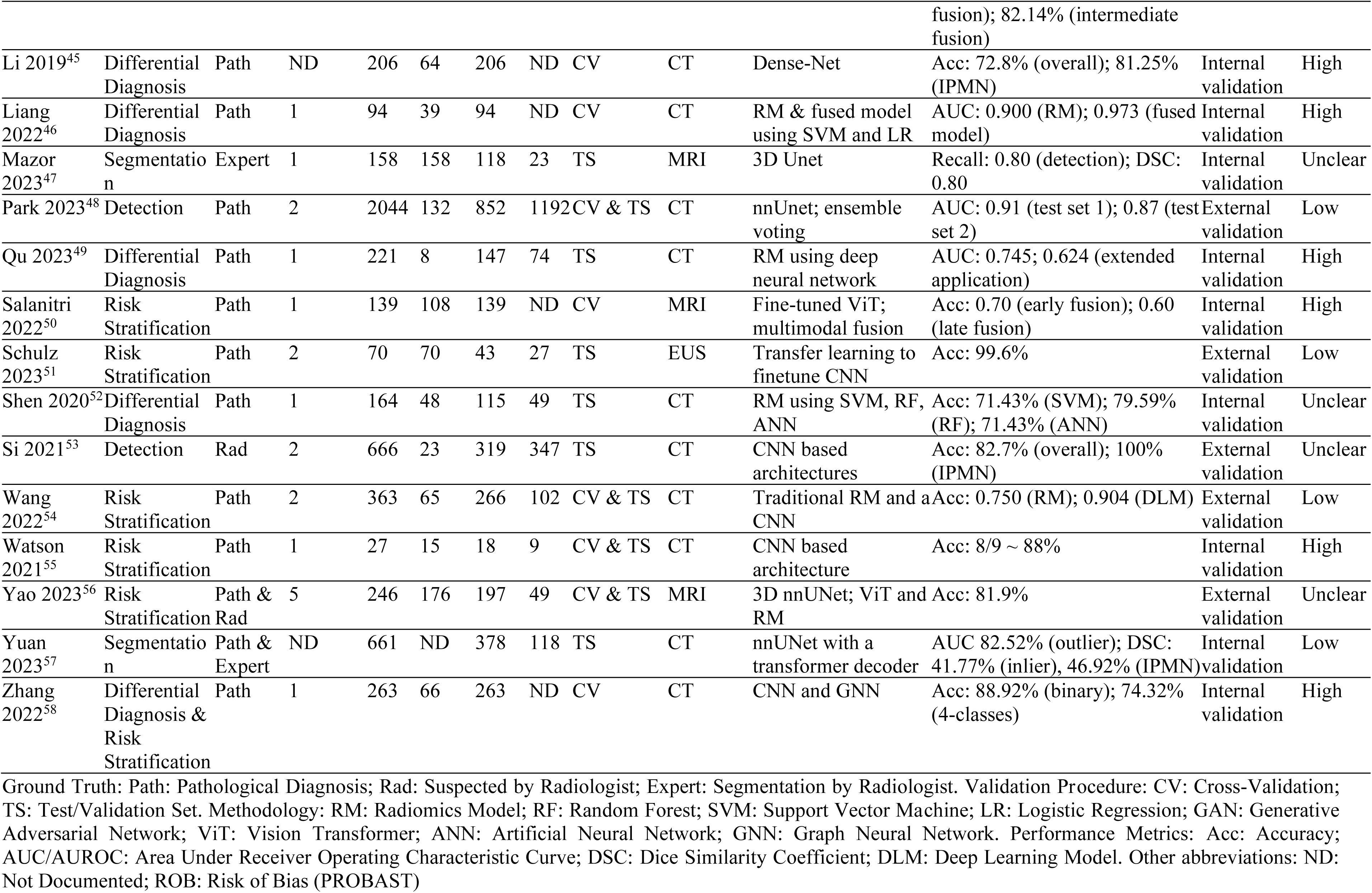
Summary and study characteristics of publications included in this systematic review.

The majority of the studies (n = 18, 72%) investigated cohorts of less than 250 patients. Three studies (12%) included over 1000 patients in total. Most of the studies had an IPMN patient population of less than 100 (n = 13, 52%), and ten studies (40%) within the range of 100-200, while only one study (4%) included over 200 patients with IPMN (Figure 3 B). Only one-third of the included studies (n = 8, 32%) mainly focused on IPMN, with the remaining studies (n = 17, 58%) including IPMN patients as a subset of total patients (Supplementary Table 2). In the studies with a large overall patient cohort size, i.e., over 1000, only a small subset of patients, under 7%, had a pathologically confirmed IPMN (Figure 3 B, Table 1). The diagnosis of IPMN or other pancreatic cysts was established through surgical pathology reports in most of the studies (n = 21, 84%), while in the other studies (n = 4, 16%) annotations provided by radiologists were considered as ground truth labels (Table 1).

The methodologies exhibit heterogeneity, yet a majority of studies used Convolutional Neural Networks (CNN) to develop the prediction model. About half of the included studies evaluate these models on independent datasets, and an almost equal number of studies present cross-validation results (Table 1).

Studies focused on IPMN/cyst detection predominantly employed the U-Net architecture for pancreas region of interest (ROI) segmentation. Alternatively, some studies used a sequential model setup with CNN-based feature extraction followed by machine learning classifiers for final detection. Reported sensitivity across these studies generally exceeded 80%. Variants of U-Net architecture optimized specifically for pancreatic cyst segmentation were commonly utilized for segmentation tasks. The reported dice similarity coefficient (DSC) for segmentation ranged from 0.42 to 0.80. For differential diagnosis, approximately half of the studies (n=4) extracted radiomic features and utilized machine learning classifiers, achieving diagnostic accuracies between 67 and 94%. The remaining studies (n=5) employed CNNs directly for image classification, with reported accuracies varying from 72 to 92%. For risk stratification, most studies implemented CNNs, with some incorporating image fusion techniques to integrate information from multiple MRI sequences. The average reported accuracy for these studies reached 85% (Figure 3 C, Table 1, Supplementary Table 2)

Risk of bias analysis using the PROBAST tool [32] identified methodological shortcomings in most included works on AI in IPMN imaging (Table 1, Supplementary Table 3).

## Discussion

Recent advances in AI for analyzing medical imaging and imaging reports have demonstrated capabilities to improve clinical tasks such as diagnosis, prognosis, therapeutic planning, and monitoring disease progression [59]. Similarly, such advanced computational methods hold promise in improving the clinical management of IPMN by enhancing diagnostic accuracy and treatment selection. We conducted a systematic review of existing publications on AI in IPMN imaging, with an emphasis on evaluating their patient and data characteristics, methodology, application areas, and stages of clinical translation.

Our analysis indicates an increasing number of publications on AI-based IPMN diagnosis and risk stratification. Nonetheless, only a limited number of studies had a primary focus on IPMN [38,41–44,50,51,56]; instead, most works included smaller numbers of IPMN patients in addition to other patient cohorts, i.e., with other types of pancreatic cysts. A prevalent challenge identified across all the publications is limited data availability, as most studies trained the AI models on small single-center datasets and did not publish data alongside the scientific analysis, resulting in an overall scarcity of high-quality PDAC datasets [60] and a notable absence of publicly available IPMN datasets. This is in contrast with the availability of datasets for other types of cancer, for instance, breast cancer [61,62] and brain tumor [63] imaging. Moreover, the reporting quality appears inadequate as several publications do not document the exact number of centers involved, which compromises the validity and reproducibility of the study [64] as well as the adoption of new technology [65]. Our risk of bias analysis using the PROBAST tool [32] mirrored such methodological shortcomings in most included works on AI in IPMN imaging. Particularly frequent sources of bias were small cohort sizes and unclear patient selection criteria.

The stratification of IPMN malignant progression based on imaging is challenging, with estimated diagnostic accuracies of imaging ranging from 60 to 80% in the scientific literature [66]. The established consensus guidelines for IPMN management and stratification recommend imaging, laboratory results, and physical examinations to stratify the risk of malignant progression [10,21]. However, the low specificity of existing methods [22] frequently leads to unnecessary surgery or surveillance of cystic lesions at high risk of malignancy. Computational analysis of pancreatic imaging can predict morphological types of IPMN noninvasively and bridge a crucial clinical gap by identifying patterns that could stratify and correlate with malignant progression in patients with IPMN. Most studies included in this systematic review proposing AI-based stratification of type of dysplasia based on imaging reported accuracies ranging from 60 to 99.6%, with most between 75 and 90% (Table 1, Supplementary Table 2).

We found CT to be the most frequently used imaging type. However, per the American College of Radiology’s Appropriateness Criteria for pancreatic cysts, the preferred imaging modality for initial evaluation is an abdominal MRI without and with intravenous contrast with magnetic resonance cholangiopancreatography (MRCP). EUS is only recommended if worrisome features are observed, and the cyst size is greater than 2.5 cm [33]. The preference for CT in literature is likely because it is more cost-effective, expeditious, and widely available in nearly all medical facilities [67], whereas MRI and EUS are protracted, costly, and comparatively less available [68]. It is noteworthy that the majority of studies rely solely on (one type of) imaging data; however, integrating multimodal clinical data could further improve model performance and enhance patient selection for surgical resection.

Methodologically, in deep learning-based studies, CNNs predominated in ROI segmentation and feature extraction, with UNet-based models, notably nnUNet [69], being the most frequently employed. In radiomics-based studies, which represented a relatively small proportion of studies investigating differential diagnosis, radiomic features (e.g., global histogram, first-order, second-order, wavelet features, etc.) are extracted and subsequently fed into classical machine learning algorithms (e.g., random forest and support vector machine) for prediction or classification. Withal, drawing definitive conclusions on the most effective AI methodology is challenging due to the dearth of large high-quality data and heterogeneity in prediction targets. While few studies directly compare different approaches on the same task or dataset, most publications report the performance of a single model on a closed- or restricted-access dataset, complicating reproducibility and comparison.

Models for language processing, particularly LLMs like GPT-4 [70] and Med-PaLM 2 [71], are increasingly being used in the field of medicine, owing to their potential to enhance patient care [72]. Despite our search strategy being inclusive of NLP and imaging reports, no such study met the inclusion criteria. However, the few retrieved studies employing language-based methods centered on the identification of patients with pancreatic cysts using rule-based algorithms [73–75]. One study used the pre-trained BioBERT model for extracting information from radiology reports in the context of pancreatic cysts, albeit without fine-tuning [76]. Therefore, the potential of modern LLMs in IPMN care, including vision-language models, remains to be elucidated. Optimization methods like Retrieval-Augmented Generation, which allows integrating authoritative knowledge such as clinical treatment guidelines in addition to its training data sources before generating outputs, may yield additional benefits over generalist LLMs in medical scenarios [77,78].

Despite various available guidelines, a standard PDAC screening examination for the general population has yet to be established. For example, routine screening mammograms are the most sensitive way to screen for breast cancer, offering relatively affordable and accessible options for screening. Multiple AI-based breast cancer screening tools have received Food and Drug Administration (FDA) approval [79] According to our review, only one study initiated a prospective clinical evaluation [36], and one study [39] extended the application to develop an end-to-end system [80] for the diagnosis of pancreatic lesions, which highlights the paucity of clinical translation and the absence of FDA-approved AI tools for screening IPMN and other pancreatic cysts.

The limitations of this systematic review are largely related to the reporting standards of the included publications. Based on the large variation in study designs, prediction targets (Table 1), and reported performance metrics, an adequate summarization of model performances across all included studies was not feasible. We therefore report a complete list of metrics (Table 1, Supplementary Table 2). We also limited our assessment of the reporting quality of included studies to a general risk of bias assessment using the PROBAST [32] tool and a detailed review of the most relevant quality criteria for AI-based studies including details on the patient cohort, methodological aspects, and the validation procedure. The reason for this was that existing tools to assess the reporting quality of studies included in systematic reviews (i.e., QUADAS [81] for systematic reviews of diagnostic accuracy studies) are not suitable for the present type of studies. Therefore, we could not perform a formal assessment of the reporting quality of the included studies.

In conclusion, this systematic review provides a comprehensive overview of the academic literature on AI in IPMN imaging and highlights the potential of computational image analysis models for the complex management of IPMN. Most existing studies rely on small, single-center patient cohorts and imaging data only. To exploit the full potential of AI for improving diagnostic accuracy and patient outcomes, it will be necessary to incorporate multicentric and multimodal clinical data and undertake robust efforts to translate retrospective research into clinical practice.

## Data Availability

All data produced in the present work are contained in the manuscript.

## Acknowledgments and Funding

The authors gratefully acknowledge assistance from Jing Lu (Purdue University, Libraries and School of Information Studies, West Lafayette, IN) in developing the search strategy for this systematic review and from Sarwesh Rauniyar (Indiana University School of Medicine, Indianapolis, IN) in developing the data extraction protocol. We thank Michaela Unger (Else Kröner Fresenius Center for Digital Health, Dresden, Germany) for providing a visualization code template for Figure 2.

CMS and FRK receive support from the Indiana Clinical and Translational Sciences Institute funded, in part, by Grant Number UM1TR004402 from the National Institutes of Health, National Center for Advancing Translational Sciences, Clinical and Translational Sciences Award. Furthermore, FRK receives support from the German Cancer Research Center (CoBot 2.0) and the Joachim Herz Foundation (Add-On Fellowship for Interdisciplinary Life Science). The content is solely the responsibility of the authors and does not necessarily represent the official views of the National Institutes of Health.

## Author Contributions

Conceptualization: MIQ, JAB, MTYS, FRK; Methodology: MIQ, JAB, MTYS, FRK; Software: MIQ, JAB, FRK; Validation: MIQ, JAB, FRK; Formal analysis: MIQ, JAB, FRK; Investigation: MIQ, JAB, MTYS, DS, TTTT, CMS, FRK; Resources: MTYS, CMS, FRK; Data Curation: MIQ, JAB, FRK; Writing - Original Draft: MIQ, FRK; Writing - Review & Editing: MIQ, JAB, MTYS, DS, TTTT, CMS, FRK; Visualization: MIQ, FRK; Supervision: MTYS, CMS, FRK; Project administration: MTYS, TTTT, CMS, FRK; Funding acquisition: MTYS, CMS, FRK.

## Supplementary Material

**Supplementary Table 1:** Data extraction template. The table presents a structured data extraction template used to extract the data from the included studies on AI in IPMN imaging.

| Parameter | Description | Example |
| --- | --- | --- |
| Year of Publication | The year in which the study was published | 2022 |
| Ground Truth Source | Source of ground truth for the prediction target | Suspected diagnosis based on radiological imaging |
| Prediction Targets | The outcomes that the proposed model aims to predict based on input data | Risk stratification of IPMN |
| Number of Centers | The total number of centers the data was collected from | 2 |
| Patient Selection Criteria | The criteria used to identify and include patients in the study. | Preoperative CT of patients with a confirmed diagnosis of IPMN. CT within 2 weeks of surgery. |
| Number of Patients the AI Model was Developed On | Number of patients included in the study, including all patients from the training, validation, and test sets. | 139 |
| Number of Patients With IPMN the AI Model was Developed On | Number of all the patients in training, validation, and test sets diagnosed with IPMN. | 110 |
| Number of Patients (Training Set) | The number of patients AI was initially trained on | 100 |
| Number of Patients (Test Set) | Number of patients AI was tested on | 39 |
| Validation Procedure | The method used to assess the performance and reliability of a model | 5-fold cross-validation or independent test set |
| Data/Imaging Modality | The type or modality of data AI model was trained on | MRI |
| Methodology | The computational methods, algorithms, and frameworks used to develop the AI model | nnUnet |
| Performance Metrics | Measures to assess the performance and accuracy of the model in predicting the outcome | Sensitivity: 95.7%, specificity: 92.6%, accuracy: 94.0% |
| Stage of Clinical Translation | Internal validation, external validation, prospective clinical evaluation, or FDA approval | External validation |

**Supplementary Table 2:**
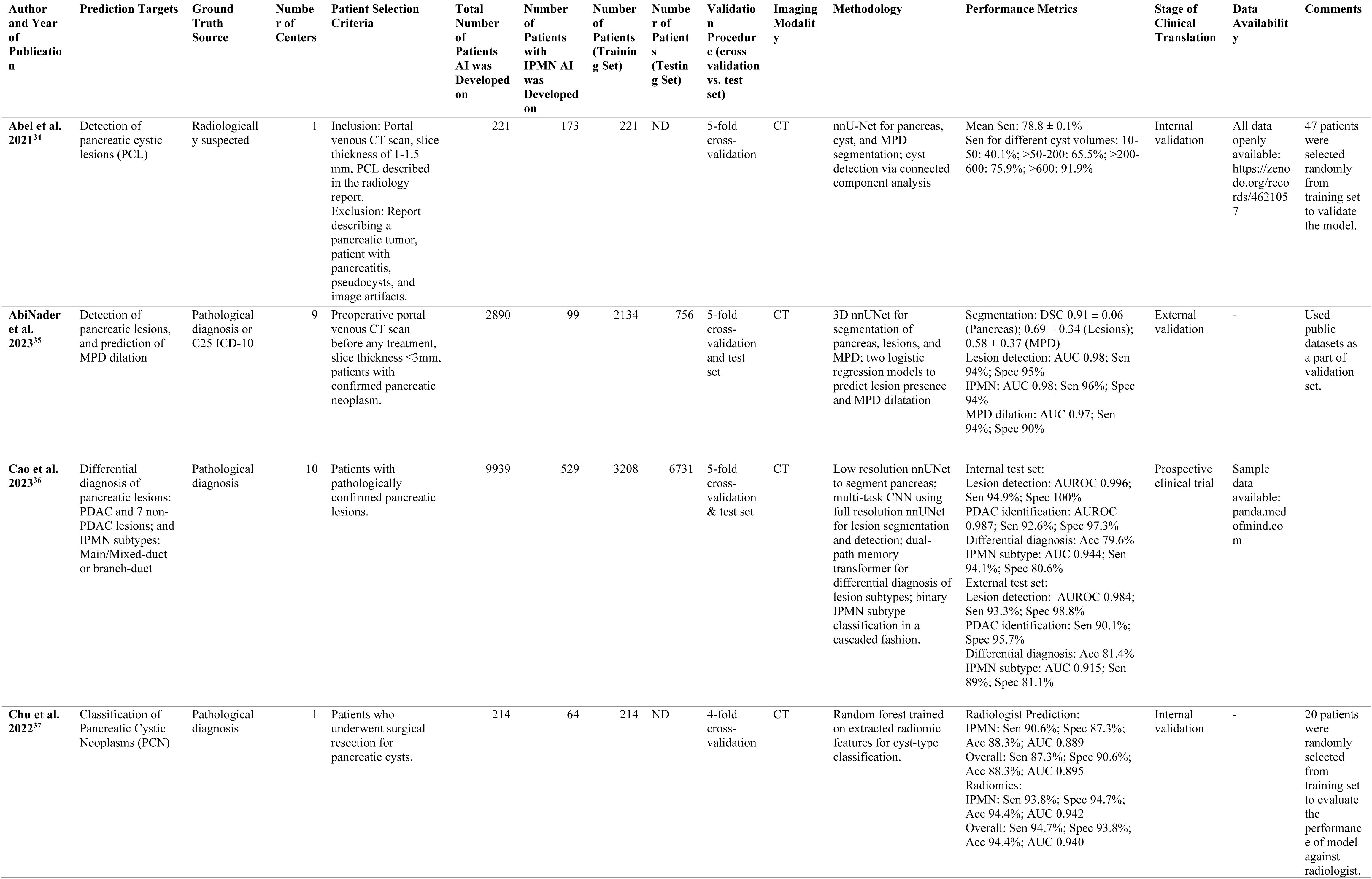
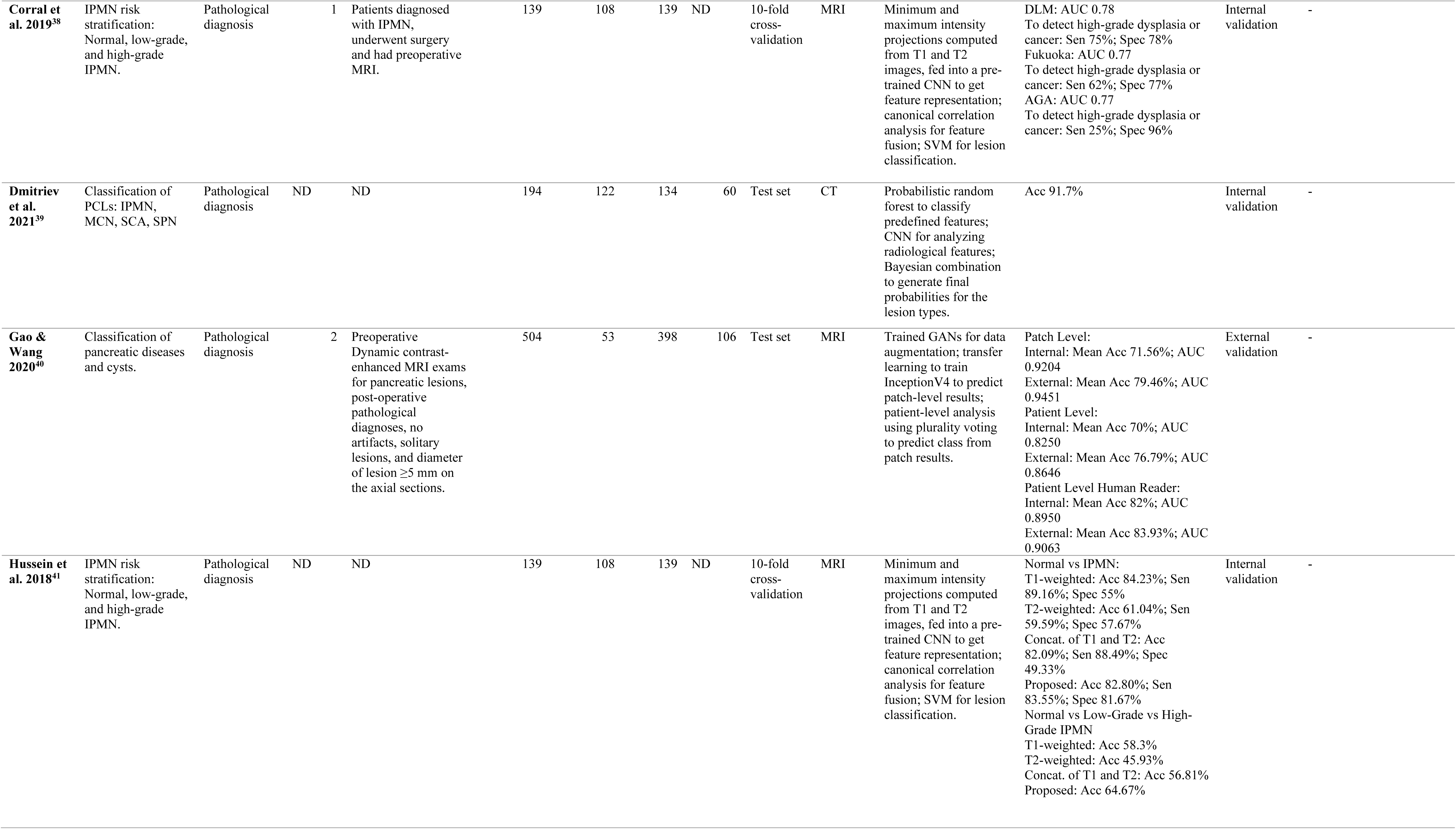
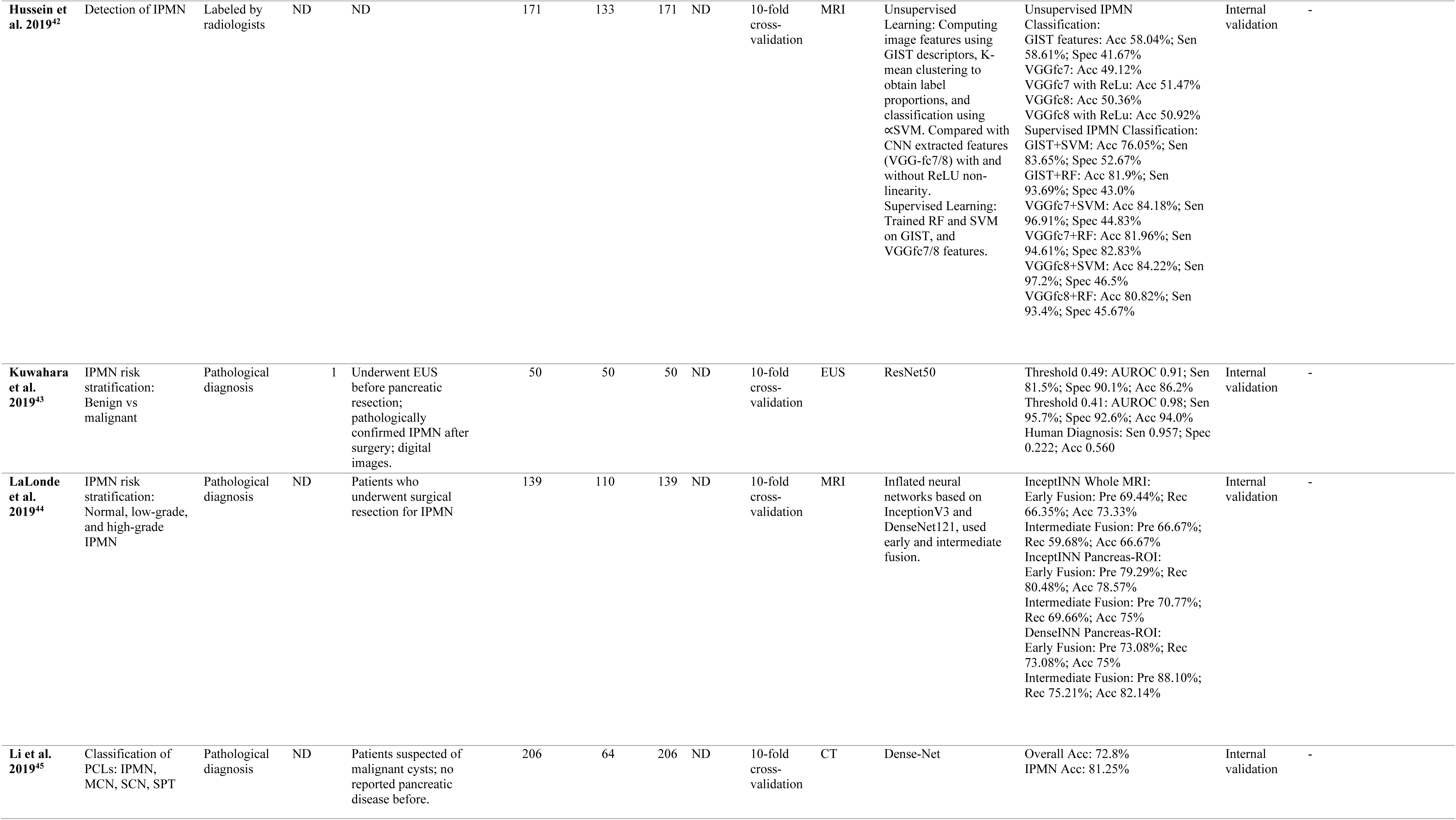
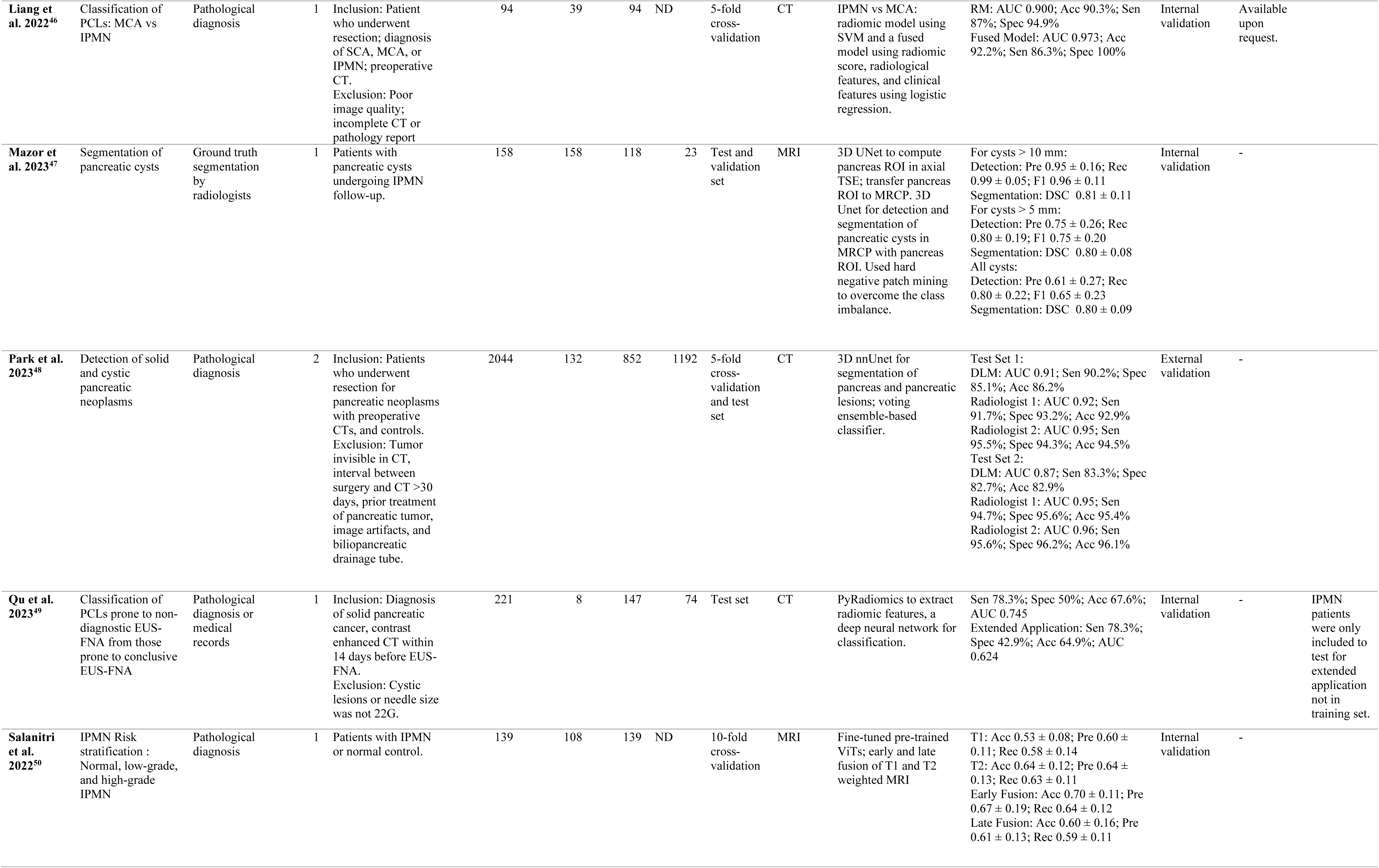
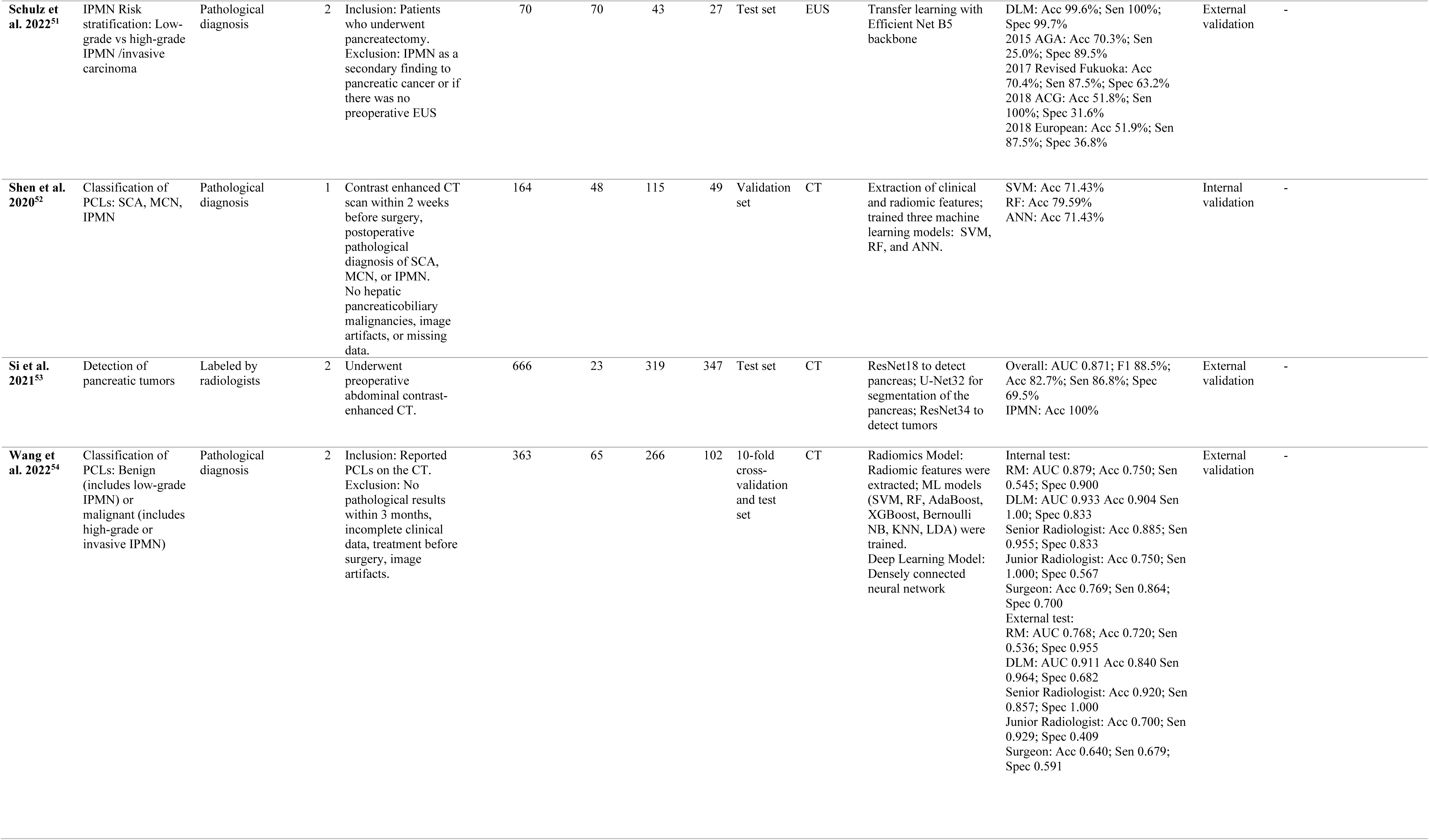
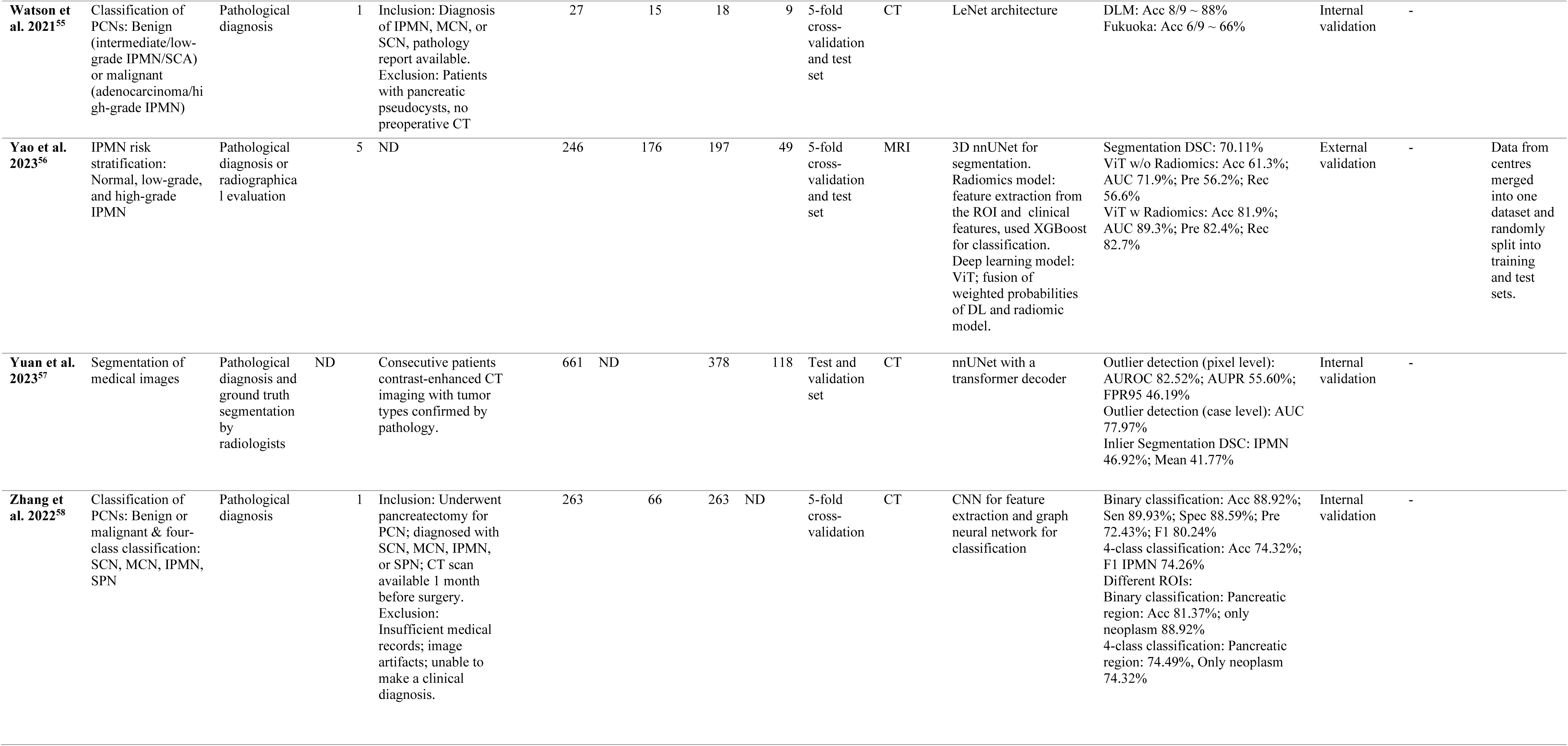
Data extracted from the included publications on AI in IPMN imaging. Abbreviations used in the table: MPD: Main Pancreatic Duct; MCN: Mucinous Cystic Neoplasm; SCA: Serous Cystadenoma; SPN: Solid Pseudopapillary Neoplasm; SCN: Serous Cystic Neoplasm; SPT: Solid Pseudopapillary Tumor; MCA: Mucinous Cystadenoma; EUS-FNA: Endoscopic Ultrasound Fine Needle Aspiration; ICD: International Classification of Diseases; ND: Not Documented; CNN: Convolutional Neural Network; SVM: Support Vector Machine; GAN: Generative Adversarial Network; RF: Random Forest; ROI: Region of Interest; ViT: Vision Transformer; ANN: Artificial Neural Network; ML: Machine Learning; NB: Naive Bayes; KNN: K-Nearest Neighbor; LDA: Linear Discriminant Analysis; DL: Deep Learning, MRCP: Magnetic Resonance Cholangiopancreatography. Performance Metrics: DLM: Deep Learning Model; RM: Radiomics Model; Sen: Sensitivity; Spec: Specificity; Acc: Accuracy; Pre: Precision; Rec: Recall; AUC/AUROC: Area Under Receiver Operating Characteristic Curve; AUPR: Area Under Precision-Recall Curve; FPR: False Positive Rate; DSC: Dice Similarity Coefficient/Dice Score.

| Author and Year of Publication | Prediction Targets | Ground Truth Source | Number of Centers | Patient Selection Criteria | Total Number of Patients AI was Developed on | Number of Patients with IPMN AI was Developed on | Number of Patients (Training Set) | Number of Patients (Testing Set) | Validation Procedure (cross validation vs. test set) | Imaging Modality | Methodology | Performance Metrics | Stage of Clinical Translation | Data Availability | Comments |
| --- | --- | --- | --- | --- | --- | --- | --- | --- | --- | --- | --- | --- | --- | --- | --- |
| Abel et al. 2021<sup>34</sup> | Detection of pancreatic cystic lesions (PCL) | Radiologically suspected | 1 | Inclusion: Portal venous CT scan, slice thickness of 1-1.5 mm, PCL described in the radiology report. Exclusion: Report describing a pancreatic tumor, patient with pancreatitis, pseudocysts, and image artifacts. | 221 | 173 | 221 | ND | 5-fold cross-validation | CT | nnU-Net for pancreas, cyst, and MPD segmentation; cyst detection via connected component analysis | Mean Sen: 78.8 ± 0.1%<br>Sen for different cyst volumes: 10-50: 40.1%; >50-200: 65.5%; >200-600: 75.9%; >600: 91.9% | Internal validation | All data openly available: https://zenodo.org/records/4621057 | 47 patients were selected randomly from training set to validate the model. |
| AbiNader et al. 2023<sup>35</sup> | Detection of pancreatic lesions, and prediction of MPD dilation | Pathological diagnosis or C25 ICD-10 | 9 | Preoperative portal venous CT scan before any treatment, slice thickness ≤3mm, patients with confirmed pancreatic neoplasm. | 2890 | 99 | 2134 | 756 | 5-fold cross-validation and test set | CT | 3D nnUNet for segmentation of pancreas, lesions, and MPD; two logistic regression models to predict lesion presence and MPD dilatation | Segmentation: DSC 0.91 ± 0.06 (Pancreas); 0.69 ± 0.34 (Lesions); 0.58 ± 0.37 (MPD)<br>Lesion detection: AUC 0.98; Sen 94%; Spec 95%<br>IPMN: AUC 0.98; Sen 96%; Spec 94%<br>MPD dilation: AUC 0.97; Sen 94%; Spec 90% | External validation | - | Used public datasets as a part of validation set. |
| Cao et al. 2023<sup>36</sup> | Differential diagnosis of pancreatic lesions: PDAC and 7 non-PDAC lesions; and IPMN subtypes: Main/Mixed-duct or branch-duct | Pathological diagnosis | 10 | Patients with pathologically confirmed pancreatic lesions. | 9939 | 529 | 3208 | 6731 | 5-fold cross-validation & test set | CT | Low resolution nnUNet to segment pancreas; multi-task CNN using full resolution nnUNet for lesion segmentation and detection; dual-path memory transformer for differential diagnosis of lesion subtypes; binary IPMN subtype classification in a cascaded fashion. | Internal test set:<br>Lesion detection: AUROC 0.996; Sen 94.9%; Spec 100%<br>PDAC identification: AUROC 0.987; Sen 92.6%; Spec 97.3%<br>Differential diagnosis: Acc 79.6%<br>IPMN subtype: AUC 0.944; Sen 94.1%; Spec 80.6%<br>External test set:<br>Lesion detection: AUROC 0.984; Sen 93.3%; Spec 98.8%<br>PDAC identification: Sen 90.1%; Spec 95.7%<br>Differential diagnosis: Acc 81.4%<br>IPMN subtype: AUC 0.915; Sen 89%; Spec 81.1% | Prospective clinical trial | Sample data available: panda.medofmind.com |  |
| Chu et al. 2022<sup>37</sup> | Classification of Pancreatic Cystic Neoplasms (PCN) | Pathological diagnosis | 1 | Patients who underwent surgical resection for pancreatic cysts. | 214 | 64 | 214 | ND | 4-fold cross-validation | CT | Random forest trained on extracted radiomic features for cyst-type classification. | Radiologist Prediction: IPMN: Sen 90.6%; Spec 87.3%; Acc 88.3%; AUC 0.889<br>Overall: Sen 87.3%; Spec 90.6%; Acc 88.3%; AUC 0.895<br>Radiomics: IPMN: Sen 93.8%; Spec 94.7%; Acc 94.4%; AUC 0.942<br>Overall: Sen 94.7%; Spec 93.8%; Acc 94.4%; AUC 0.940 | Internal validation | - | 20 patients were randomly selected from training set to evaluate the performance of model against radiologist. |

|  |  |  |  |  |  |  |  |  |  |  |  |  |  |  |
| --- | --- | --- | --- | --- | --- | --- | --- | --- | --- | --- | --- | --- | --- | --- |
| **Corral et al. 2019**<sup>38</sup> | IPMN risk stratification: Normal, low-grade, and high-grade IPMN. | Pathological diagnosis | 1 | Patients diagnosed with IPMN, underwent surgery and had preoperative MRI. | 139 | 108 | 139 | ND | 10-fold cross-validation | MRI | Minimum and maximum intensity projections computed from T1 and T2 images, fed into a pre-trained CNN to get feature representation; canonical correlation analysis for feature fusion; SVM for lesion classification. | DLM: AUC 0.78<br>To detect high-grade dysplasia or cancer: Sen 75%; Spec 78%<br>Fukuoka: AUC 0.77<br>To detect high-grade dysplasia or cancer: Sen 62%; Spec 77%<br>AGA: AUC 0.77<br>To detect high-grade dysplasia or cancer: Sen 25%; Spec 96% | Internal validation | - |
| **Dmitriev et al. 2021**<sup>39</sup> | Classification of PCLs: IPMN, MCN, SCA, SPN | Pathological diagnosis | ND | ND | 194 | 122 | 134 | 60 | Test set | CT | Probabilistic random forest to classify predefined features; CNN for analyzing radiological features; Bayesian combination to generate final probabilities for the lesion types. | Acc 91.7% | Internal validation | - |
| **Gao & Wang 2020**<sup>40</sup> | Classification of pancreatic diseases and cysts. | Pathological diagnosis | 2 | Preoperative Dynamic contrast-enhanced MRI exams for pancreatic lesions, post-operative pathological diagnoses, no artifacts, solitary lesions, and diameter of lesion $\geq 5$ mm on the axial sections. | 504 | 53 | 398 | 106 | Test set | MRI | Trained GANs for data augmentation; transfer learning to train InceptionV4 to predict patch-level results; patient-level analysis using plurality voting to predict class from patch results. | Patch Level:<br>Internal: Mean Acc 71.56%; AUC 0.9204<br>External: Mean Acc 79.46%; AUC 0.9451<br>Patient Level:<br>Internal: Mean Acc 70%; AUC 0.8250<br>External: Mean Acc 76.79%; AUC 0.8646<br>Patient Level Human Reader:<br>Internal: Mean Acc 82%; AUC 0.8950<br>External: Mean Acc 83.93%; AUC 0.9063 | External validation | - |
| **Hussein et al. 2018**<sup>41</sup> | IPMN risk stratification: Normal, low-grade, and high-grade IPMN. | Pathological diagnosis | ND | ND | 139 | 108 | 139 | ND | 10-fold cross-validation | MRI | Minimum and maximum intensity projections computed from T1 and T2 images, fed into a pre-trained CNN to get feature representation; canonical correlation analysis for feature fusion; SVM for lesion classification. | Normal vs IPMN:<br>T1-weighted: Acc 84.23%; Sen 89.16%; Spec 55%<br>T2-weighted: Acc 61.04%; Sen 59.59%; Spec 57.67%<br>Concat. of T1 and T2: Acc 82.09%; Sen 88.49%; Spec 49.33%<br>Proposed: Acc 82.80%; Sen 83.55%; Spec 81.67%<br>Normal vs Low-Grade vs High-Grade IPMN<br>T1-weighted: Acc 58.3%<br>T2-weighted: Acc 45.93%<br>Concat. of T1 and T2: Acc 56.81%<br>Proposed: Acc 64.67% | Internal validation | - |
| **Hussein et al. 2019**<sup>42</sup> | Detection of IPMN | Labeled by radiologists | ND | ND | 171 | 133 | 171 | ND | 10-fold cross-validation | MRI | Unsupervised Learning: Computing image features using GIST descriptors, K-mean clustering to obtain label proportions, and classification using αSVM. Compared with CNN extracted features (VGG-fc7/8) with and without ReLU non-linearity. Supervised Learning: Trained RF and SVM on GIST, and VGGfc7/8 features. | Unsupervised IPMN Classification: GIST features: Acc 58.04%; Sen 58.61%; Spec 41.67%<br>VGGfc7: Acc 49.12%<br>VGGfc7 with ReLu: Acc 51.47%<br>VGGfc8: Acc 50.36%<br>VGGfc8 with ReLu: Acc 50.92%<br>Supervised IPMN Classification: GIST+SVM: Acc 76.05%; Sen 83.65%; Spec 52.67%<br>GIST+RF: Acc 81.9%; Sen 93.69%; Spec 43.0%<br>VGGfc7+SVM: Acc 84.18%; Sen 96.91%; Spec 44.83%<br>VGGfc7+RF: Acc 81.96%; Sen 94.61%; Spec 82.83%<br>VGGfc8+SVM: Acc 84.22%; Sen 97.2%; Spec 46.5%<br>VGGfc8+RF: Acc 80.82%; Sen 93.4%; Spec 45.67% | Internal validation | - |
| **Kuwahara et al. 2019**<sup>43</sup> | IPMN risk stratification: Benign vs malignant | Pathological diagnosis | 1 | Underwent EUS before pancreatic resection; pathologically confirmed IPMN after surgery; digital images. | 50 | 50 | 50 | ND | 10-fold cross-validation | EUS | ResNet50 | Threshold 0.49: AUROC 0.91; Sen 81.5%; Spec 90.1%; Acc 86.2%<br>Threshold 0.41: AUROC 0.98; Sen 95.7%; Spec 92.6%; Acc 94.0%<br>Human Diagnosis: Sen 0.957; Spec 0.222; Acc 0.560 | Internal validation | - |
| **LaLonde et al. 2019**<sup>44</sup> | IPMN risk stratification: Normal, low-grade, and high-grade IPMN | Pathological diagnosis | ND | Patients who underwent surgical resection for IPMN | 139 | 110 | 139 | ND | 10-fold cross-validation | MRI | Inflated neural networks based on InceptionV3 and DenseNet121, used early and intermediate fusion. | InceptINN Whole MRI: Early Fusion: Pre 69.44%; Rec 66.35%; Acc 73.33%<br>Intermediate Fusion: Pre 66.67%; Rec 59.68%; Acc 66.67%<br>InceptINN Pancreas-ROI: Early Fusion: Pre 79.29%; Rec 80.48%; Acc 78.57%<br>Intermediate Fusion: Pre 70.77%; Rec 69.66%; Acc 75%<br>DenseINN Pancreas-ROI: Early Fusion: Pre 73.08%; Rec 73.08%; Acc 75%<br>Intermediate Fusion: Pre 88.10%; Rec 75.21%; Acc 82.14% | Internal validation | - |
| **Li et al. 2019**<sup>45</sup> | Classification of PCLs: IPMN, MCN, SCN, SPT | Pathological diagnosis | ND | Patients suspected of malignant cysts; no reported pancreatic disease before. | 206 | 64 | 206 | ND | 10-fold cross-validation | CT | Dense-Net | Overall Acc: 72.8%<br>IPMN Acc: 81.25% | Internal validation | - |
| **Liang et al. 2022**<sup>46</sup> | Classification of PCLs: MCA vs IPMN | Pathological diagnosis | 1 | Inclusion: Patient who underwent resection; diagnosis of SCA, MCA, or IPMN; preoperative CT.<br>Exclusion: Poor image quality; incomplete CT or pathology report | 94 | 39 | 94 | ND | 5-fold cross-validation | CT | IPMN vs MCA: radiomic model using SVM and a fused model using radiomic score, radiological features, and clinical features using logistic regression. | RM: AUC 0.900; Acc 90.3%; Sen 87%; Spec 94.9%<br>Fused Model: AUC 0.973; Acc 92.2%; Sen 86.3%; Spec 100% | Internal validation | Available upon request. |  |
| **Mazor et al. 2023**<sup>47</sup> | Segmentation of pancreatic cysts | Ground truth segmentation by radiologists | 1 | Patients with pancreatic cysts undergoing IPMN follow-up. | 158 | 158 | 118 | 23 | Test and validation set | MRI | 3D UNet to compute pancreas ROI in axial TSE; transfer pancreas ROI to MRCP. 3D Unet for detection and segmentation of pancreatic cysts in MRCP with pancreas ROI. Used hard negative patch mining to overcome the class imbalance. | For cysts > 10 mm:<br>Detection: Pre $0.95 \pm 0.16$; Rec $0.99 \pm 0.05$; F1 $0.96 \pm 0.11$<br>Segmentation: DSC $0.81 \pm 0.11$<br>For cysts > 5 mm:<br>Detection: Pre $0.75 \pm 0.26$; Rec $0.80 \pm 0.19$; F1 $0.75 \pm 0.20$<br>Segmentation: DSC $0.80 \pm 0.08$<br>All cysts:<br>Detection: Pre $0.61 \pm 0.27$; Rec $0.80 \pm 0.22$; F1 $0.65 \pm 0.23$<br>Segmentation: DSC $0.80 \pm 0.09$ | Internal validation | - | |
| **Park et al. 2023**<sup>48</sup> | Detection of solid and cystic pancreatic neoplasms | Pathological diagnosis | 2 | Inclusion: Patients who underwent resection for pancreatic neoplasms with preoperative CTs, and controls.<br>Exclusion: Tumor invisible in CT, interval between surgery and CT >30 days, prior treatment of pancreatic tumor, image artifacts, and biliopancreatic drainage tube. | 2044 | 132 | 852 | 1192 | 5-fold cross-validation and test set | CT | 3D nnUnet for segmentation of pancreas and pancreatic lesions; voting ensemble-based classifier. | Test Set 1:<br>DLM: AUC 0.91; Sen 90.2%; Spec 85.1%; Acc 86.2%<br>Radiologist 1: AUC 0.92; Sen 91.7%; Spec 93.2%; Acc 92.9%<br>Radiologist 2: AUC 0.95; Sen 95.5%; Spec 94.3%; Acc 94.5%<br>Test Set 2:<br>DLM: AUC 0.87; Sen 83.3%; Spec 82.7%; Acc 82.9%<br>Radiologist 1: AUC 0.95; Sen 94.7%; Spec 95.6%; Acc 95.4%<br>Radiologist 2: AUC 0.96; Sen 95.6%; Spec 96.2%; Acc 96.1% | External validation | - |  |
| **Qu et al. 2023**<sup>49</sup> | Classification of PCLs prone to non-diagnostic EUS-FNA from those prone to conclusive EUS-FNA | Pathological diagnosis or medical records | 1 | Inclusion: Diagnosis of solid pancreatic cancer, contrast enhanced CT within 14 days before EUS-FNA.<br>Exclusion: Cystic lesions or needle size was not 22G. | 221 | 8 | 147 | 74 | Test set | CT | PyRadiomics to extract radiomic features, a deep neural network for classification. | Sen 78.3%; Spec 50%; Acc 67.6%; AUC 0.745<br>Extended Application: Sen 78.3%; Spec 42.9%; Acc 64.9%; AUC 0.624 | Internal validation | - | IPMN patients were only included to test for extended application not in training set. |
| **Salanitri et al. 2022**<sup>50</sup> | IPMN Risk stratification : Normal, low-grade, and high-grade IPMN | Pathological diagnosis | 1 | Patients with IPMN or normal control. | 139 | 108 | 139 | ND | 10-fold cross-validation | MRI | Fine-tuned pre-trained ViTs; early and late fusion of T1 and T2 weighted MRI | T1: Acc $0.53 \pm 0.08$; Pre $0.60 \pm 0.11$; Rec $0.58 \pm 0.14$<br>T2: Acc $0.64 \pm 0.12$; Pre $0.64 \pm 0.13$; Rec $0.63 \pm 0.11$<br>Early Fusion: Acc $0.70 \pm 0.11$; Pre $0.67 \pm 0.19$; Rec $0.64 \pm 0.12$<br>Late Fusion: Acc $0.60 \pm 0.16$; Pre $0.61 \pm 0.13$; Rec $0.59 \pm 0.11$ | Internal validation | - | |

|  |  |  |  |  |  |  |  |  |  |  |  |  |  |  |
| --- | --- | --- | --- | --- | --- | --- | --- | --- | --- | --- | --- | --- | --- | --- |
| **Schulz et al. 2022**<sup>51</sup> | IPMN Risk stratification: Low-grade vs high-grade IPMN /invasive carcinoma | Pathological diagnosis | 2 | Inclusion: Patients who underwent pancreatectomy. Exclusion: IPMN as a secondary finding to pancreatic cancer or if there was no preoperative EUS | 70 | 70 | 43 | 27 | Test set | EUS | Transfer learning with Efficient Net B5 backbone | DLM: Acc 99.6%; Sen 100%; Spec 99.7%<br>2015 AGA: Acc 70.3%; Sen 25.0%; Spec 89.5%<br>2017 Revised Fukuoka: Acc 70.4%; Sen 87.5%; Spec 63.2%<br>2018 ACG: Acc 51.8%; Sen 100%; Spec 31.6%<br>2018 European: Acc 51.9%; Sen 87.5%; Spec 36.8% | External validation | - |
| **Shen et al. 2020**<sup>52</sup> | Classification of PCLs: SCA, MCN, IPMN | Pathological diagnosis | 1 | Contrast enhanced CT scan within 2 weeks before surgery, postoperative pathological diagnosis of SCA, MCN, or IPMN. No hepatic pancreaticobiliary malignancies, image artifacts, or missing data. | 164 | 48 | 115 | 49 | Validation set | CT | Extraction of clinical and radiomic features; trained three machine learning models: SVM, RF, and ANN. | SVM: Acc 71.43%<br>RF: Acc 79.59%<br>ANN: Acc 71.43% | Internal validation | - |
| **Si et al. 2021**<sup>53</sup> | Detection of pancreatic tumors | Labeled by radiologists | 2 | Underwent preoperative abdominal contrast-enhanced CT. | 666 | 23 | 319 | 347 | Test set | CT | ResNet18 to detect pancreas; U-Net32 for segmentation of the pancreas; ResNet34 to detect tumors | Overall: AUC 0.871; F1 88.5%; Acc 82.7%; Sen 86.8%; Spec 69.5%<br>IPMN: Acc 100% | External validation | - |
| **Wang et al. 2022**<sup>54</sup> | Classification of PCLs: Benign (includes low-grade IPMN) or malignant (includes high-grade or invasive IPMN) | Pathological diagnosis | 2 | Inclusion: Reported PCLs on the CT. Exclusion: No pathological results within 3 months, incomplete clinical data, treatment before surgery, image artifacts. | 363 | 65 | 266 | 102 | 10-fold cross-validation and test set | CT | Radiomics Model: Radiomic features were extracted; ML models (SVM, RF, AdaBoost, XGBoost, Bernoulli NB, KNN, LDA) were trained. Deep Learning Model: Densely connected neural network | Internal test:<br>RM: AUC 0.879; Acc 0.750; Sen 0.545; Spec 0.900<br>DLM: AUC 0.933 Acc 0.904 Sen 1.00; Spec 0.833<br>Senior Radiologist: Acc 0.885; Sen 0.955; Spec 0.833<br>Junior Radiologist: Acc 0.750; Sen 1.000; Spec 0.567<br>Surgeon: Acc 0.769; Sen 0.864; Spec 0.700<br>External test:<br>RM: AUC 0.768; Acc 0.720; Sen 0.536; Spec 0.955<br>DLM: AUC 0.911 Acc 0.840 Sen 0.964; Spec 0.682<br>Senior Radiologist: Acc 0.920; Sen 0.857; Spec 1.000<br>Junior Radiologist: Acc 0.700; Sen 0.929; Spec 0.409<br>Surgeon: Acc 0.640; Sen 0.679; Spec 0.591 | External validation | - |
| **Watson et al. 2021**<sup>55</sup> | Classification of PCNs: Benign (intermediate/low-grade IPMN/SCA) or malignant (adenocarcinoma/high-grade IPMN) | Pathological diagnosis | 1 | Inclusion: Diagnosis of IPMN, MCN, or SCN, pathology report available. Exclusion: Patients with pancreatic pseudocysts, no preoperative CT | 27 | 15 | 18 | 9 | 5-fold cross-validation and test set | CT | LeNet architecture | DLM: Acc 8/9 ~ 88%<br>Fukuoka: Acc 6/9 ~ 66% | Internal validation | - |  |
| **Yao et al. 2023**<sup>56</sup> | IPMN risk stratification: Normal, low-grade, and high-grade IPMN | Pathological diagnosis or radiographical evaluation | 5 | ND | 246 | 176 | 197 | 49 | 5-fold cross-validation and test set | MRI | 3D nnUNet for segmentation. Radiomics model: feature extraction from the ROI and clinical features, used XGBoost for classification. Deep learning model: ViT; fusion of weighted probabilities of DL and radiomic model. | Segmentation DSC: 70.11%<br>ViT w/o Radiomics: Acc 61.3%; AUC 71.9%; Pre 56.2%; Rec 56.6%<br>ViT w Radiomics: Acc 81.9%; AUC 89.3%; Pre 82.4%; Rec 82.7% | External validation | - | Data from centres merged into one dataset and randomly split into training and test sets. |
| **Yuan et al. 2023**<sup>57</sup> | Segmentation of medical images | Pathological diagnosis and ground truth segmentation by radiologists | ND | Consecutive patients contrast-enhanced CT imaging with tumor types confirmed by pathology. | 661 | ND | 378 | 118 | Test and validation set | CT | nnUNet with a transformer decoder | Outlier detection (pixel level): AUROC 82.52%; AUPR 55.60%; FPR95 46.19%<br>Outlier detection (case level): AUC 77.97%<br>Inlier Segmentation DSC: IPMN 46.92%; Mean 41.77% | Internal validation | - |  |
| **Zhang et al. 2022**<sup>58</sup> | Classification of PCNs: Benign or malignant & four-class classification: SCN, MCN, IPMN, SPN | Pathological diagnosis | 1 | Inclusion: Underwent pancreatectomy for PCN; diagnosed with SCN, MCN, IPMN, or SPN; CT scan available 1 month before surgery. Exclusion: Insufficient medical records; image artifacts; unable to make a clinical diagnosis. | 263 | 66 | 263 | ND | 5-fold cross-validation | CT | CNN for feature extraction and graph neural network for classification | Binary classification: Acc 88.92%; Sen 89.93%; Spec 88.59%; Pre 72.43%; F1 80.24%<br>4-class classification: Acc 74.32%; F1 IPMN 74.26%<br>Different ROIs:<br>Binary classification: Pancreatic region: Acc 81.37%; only neoplasm 88.92%<br>4-class classification: Pancreatic region: 74.49%, Only neoplasm 74.32% | Internal validation | - |  |

**Supplementary Table 3:** Risk of bias assessment using PROBAST Tool.

| **Study ID** | **Participants** | **Predictor** | **Outcome** | **Analysis** | **Overall** |
| --- | --- | --- | --- | --- | --- |
| **Abel 2021** | Low | Low | Unclear | Low | High |
| **AbiNader**<br>**2023** | Low | Low | Unclear | Low | Low |
| **Cao 2023** | Low | Low | Low | Low | Low |
| **Chu 2022** | Unclear | Low | Low | Unclear | High |
| **Corral 2019** | Low | Low | Low | Unclear | High |
| **Dmitriev 2021** | High | Low | Low | High | High |
| **Gao 2020** | Low | Low | Low | Low | Low |
| **Hussein 2017** | High | Low | Low | Unclear | High |
| **Hussein 2019** | High | Low | High | Low | High |
| **Kuwahara**<br>**2019** | Low | Low | Low | Unclear | High |
| **LaLonde 2019** | High | Low | Low | Unclear | High |
| **Li 2019** | Unclear | Low | Low | Unclear | High |
| **Liang 2022** | Low | Low | Low | Low | High |
| **Mazor 2023** | Unclear | Low | Low | Low | Unclear |
| **Park 2023** | Low | Low | Low | Low | Low |
| **Qu 2023** | Low | Low | Unclear | High | High |
| **Salanitri 2022** | Unclear | Low | Low | Low | High |
| **Schulz 2022** | Low | Low | Low | Low | Low |
| **Shen 2020** | Low | Low | Low | Unclear | Unclear |
| **Si 2021** | Unclear | Low | Unclear | Low | Unclear |
| **Wang 2022** | Low | Low | Low | Low | Low |
| **Watson 2021** | Low | Low | Low | High | High |
| **Yao 2023** | Unclear | Low | Unclear | Low | Unclear |
| **Yuan 2023** | Low | Low | Low | Low | Low |
| **Zhang 2022** | Low | Low | Low | Low | High |

**Supplementary Figure 1:**
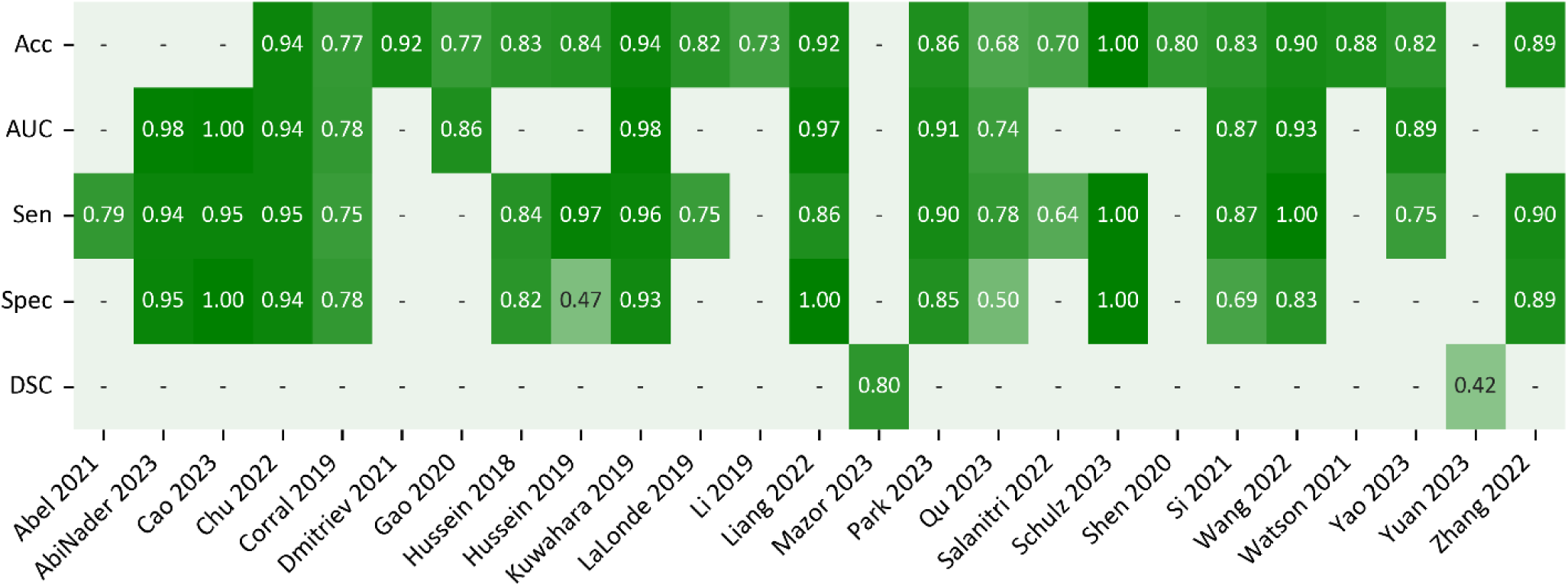
Performance Metrics Across Prediction Targets. For detection, differential diagnosis, and risk stratification accuracy (Acc), area under the receiver operating curve (AUC), sensitivity (Sen), and specificity (Spec) are presented. For segmentation, the DSC is presented. — indicates that the metric was not reported.

## PRISMA 2020 Checklist

| Section and Topic | Item # | Checklist item | Location where item is reported |
| --- | --- | --- | --- |
| **TITLE** |  |  |  |
| Title | 1 | Identify the report as a systematic review. | Manuscript – Page 1 |
| **ABSTRACT** |  |  |  |
| Abstract | 2 | See the PRISMA 2020 for Abstracts checklist. | Main Body – Page 1 |
| **INTRODUCTION** |  |  |  |
| Rationale | 3 | Describe the rationale for the review in the context of existing knowledge. | Main Body – Page 3,4 |
| Objectives | 4 | Provide an explicit statement of the objective(s) or question(s) the review addresses. | Main Body – Page 3,4 |
| **METHODS** |  |  |  |
| Eligibility criteria | 5 | Specify the inclusion and exclusion criteria for the review and how studies were grouped for the syntheses. | Main Body – Page 4,5 |
| Information sources | 6 | Specify all databases, registers, websites, organisations, reference lists and other sources searched or consulted to identify studies. Specify the date when each source was last searched or consulted. | Main Body – Page 4,5 |
| Search strategy | 7 | Present the full search strategies for all databases, registers and websites, including any filters and limits used. | Main Body – Page 4,5 |
| Selection process | 8 | Specify the methods used to decide whether a study met the inclusion criteria of the review, including how many reviewers screened each record and each report retrieved, whether they worked independently, and if applicable, details of automation tools used in the process. | Main Body – Page 4,5 |
| Data collection process | 9 | Specify the methods used to collect data from reports, including how many reviewers collected data from each report, whether they worked independently, any processes for obtaining or confirming data from study investigators, and if applicable, details of automation tools used in the process. | Main Body – Page 4,5 |
| Data items | 10a | List and define all outcomes for which data were sought. Specify whether all results that were compatible with each outcome domain in each study were sought (e.g. for all measures, time points, analyses), and if not, the methods used to decide which results to collect. | Main Body – Page 4,5 |
|  | 10b | List and define all other variables for which data were sought (e.g. participant and intervention characteristics, funding sources). Describe any assumptions made about any missing or unclear information. | Main Body – Page 5,6,7 |
| Study risk of bias assessment | 11 | Specify the methods used to assess risk of bias in the included studies, including details of the tool(s) used, how many reviewers assessed each study and whether they worked independently, and if applicable, details of automation tools used in the process. | Main Body – Page 5 |
| Effect measures | 12 | Specify for each outcome the effect measure(s) (e.g. risk ratio, mean difference) used in the synthesis or presentation of results. | Main Body – Page 5 |
| Synthesis methods | 13a | Describe the processes used to decide which studies were eligible for each synthesis (e.g. tabulating the study intervention characteristics and comparing against the planned groups for each synthesis (item #5)). | Main Body – Page 4,5 |
|  | 13b | Describe any methods required to prepare the data for presentation or synthesis, such as handling of missing summary statistics, or data conversions. | Main Body – Page 4,5 |
|  | 13c | Describe any methods used to tabulate or visually display results of individual studies and syntheses. | Main Body – |

PRISMA 2020 Checklist
| Section and Topic | Item # | Checklist item | Location where item is reported |
| --- | --- | --- | --- |
|  |  |  | Page 4,5 |
|  | 13d | Describe any methods used to synthesize results and provide a rationale for the choice(s). If meta-analysis was performed, describe the model(s), method(s) to identify the presence and extent of statistical heterogeneity, and software package(s) used. | Main Body – Page 4,5 |
|  | 13e | Describe any methods used to explore possible causes of heterogeneity among study results (e.g. subgroup analysis, meta-regression). | Main Body – Page 4,5 |
|  | 13f | Describe any sensitivity analyses conducted to assess robustness of the synthesized results. | Main Body – Page 4,5 |
| Reporting bias assessment | 14 | Describe any methods used to assess risk of bias due to missing results in a synthesis (arising from reporting biases). | Main Body – Page 4,5 |
| Certainty assessment | 15 | Describe any methods used to assess certainty (or confidence) in the body of evidence for an outcome. | Main Body – Page 4,5 |
| **RESULTS** |  |  |  |
| Study selection | 16a | Describe the results of the search and selection process, from the number of records identified in the search to the number of studies included in the review, ideally using a flow diagram. | Main Body – Page 5-9<br>Figure 1 |
|  | 16b | Cite studies that might appear to meet the inclusion criteria, but which were excluded, and explain why they were excluded. | Main Body – Page 5-9 |
| Study characteristics | 17 | Cite each included study and present its characteristics. | Table 1,<br>Supplementary Table 2 |
| Risk of bias in studies | 18 | Present assessments of risk of bias for each included study. | Table 1,<br>Supplementary Table 3 |
| Results of individual studies | 19 | For all outcomes, present, for each study: (a) summary statistics for each group (where appropriate) and (b) an effect estimate and its precision (e.g. confidence/credible interval), ideally using structured tables or plots. | Table 1, Figure 2,3 |
| Results of syntheses | 20a | For each synthesis, briefly summarise the characteristics and risk of bias among contributing studies. | Main Body – Page 6-9<br>Table 1,<br>Supplementary Table 3 |
|  | 20b | Present results of all statistical syntheses conducted. If meta-analysis was done, present for each the summary estimate and its precision (e.g. confidence/credible interval) and measures of statistical heterogeneity. If comparing groups, describe the direction of the effect. | Main Body – Page 5-9 |
|  | 20c | Present results of all investigations of possible causes of heterogeneity among study results. | Main Body – Page 5-9 |
|  | 20d | Present results of all sensitivity analyses conducted to assess the robustness of the synthesized results. | Main Body – Page 5-9 |
| Reporting biases | 21 | Present assessments of risk of bias due to missing results (arising from reporting biases) for each synthesis assessed. | Main Body – |

PRISMA 2020 Checklist
| Section and Topic | Item # | Checklist item | Location where item is reported |
| --- | --- | --- | --- |
|  |  |  | Page 5-9<br>Table 1,<br>Supplementary<br>Table 3 |
| Certainty of evidence | 22 | Present assessments of certainty (or confidence) in the body of evidence for each outcome assessed. | Main Body –<br>Page 5-9 |
| **DISCUSSION** |  |  |  |
| Discussion | 23a | Provide a general interpretation of the results in the context of other evidence. | Main Body –<br>Page 12-14 |
|  | 23b | Discuss any limitations of the evidence included in the review. | Main Body –<br>Page 12-14 |
|  | 23c | Discuss any limitations of the review processes used. | Main Body –<br>Page 12-14 |
|  | 23d | Discuss implications of the results for practice, policy, and future research. | Main Body –<br>Page 12-14 |
| **OTHER INFORMATION** |  |  |  |
| Registration and protocol | 24a | Provide registration information for the review, including register name and registration number, or state that the review was not registered. | Main Body –<br>Page 5 |
|  | 24b | Indicate where the review protocol can be accessed, or state that a protocol was not prepared. | Main Body –<br>Page 5 |
|  | 24c | Describe and explain any amendments to information provided at registration or in the protocol. | Main Body –<br>Page 5 |
| Support | 25 | Describe sources of financial or non-financial support for the review, and the role of the funders or sponsors in the review. | Main Body –<br>15 |
| Competing interests | 26 | Declare any competing interests of review authors. | Main Body –<br>15 |
| Availability of data, code and other materials | 27 | Report which of the following are publicly available and where they can be found: template data collection forms; data extracted from included studies; data used for all analyses; analytic code; any other materials used in the review. | Supplementary<br>Table 1,2,3 |

From: Page MJ, McKenzie JE, Bossuyt PM, Boutron I, Hoffmann TC, Mulrow CD, et al. The PRISMA 2020 statement: an updated guideline for reporting systematic reviews. BMJ 2021;372:n71. doi: 10.1136/bmj.n71

